# Advancing Brain Tumor Diagnosis Using Deep Learning: A Systematic Review on Glioma Segmentation and Classification on Multiparametric MRI

**DOI:** 10.64898/2026.01.13.26344038

**Authors:** Simona Aresta, Cinzia Palmirotta, Muhammad Asim, Petronilla Battista, Claudia Cava, Pietro Fiore, Andrea Santamato, Paolo Vitali, Isabella Castiglioni, Gennaro D’Anna, Leonardo Rundo, Christian Salvatore

## Abstract

Brain tumors are among the most lethal cancers with gliomas representing the most morphologically complex type. Precise and time efficient glioma segmentation and classification are essential for accurate diagnosis, treatment planning, and patient monitoring. Magnetic resonance imaging (MRI) remains the primary imaging modality for noninvasive glioma assessment. This review systematically analyzes deep learning (DL) and artificial intelligence (AI) approaches for brain tumor segmentation and classification. Thirty one studies, out of 310 published between 2022 and 2025, met the inclusion criteria, among which 8 performed both segmentation and classification tasks. For segmentation, most of the studies used publicly available multiparametric MRI datasets. Segmentation performance varied by model and tumor region, with those focused on the whole tumor region achieving the highest Dice Score Coefficient (DSC). Classical U Nets achieved DSC scores around 80%, while advanced models integrating residual or attention modules exceeded 90%. Two main classification tasks were performed: tumor type and glioma staging. Classification models primarily relied on learned features extracted from multiparametric MRI using DL models, reporting an accuracy from 91.3% to 99.4%, with sensitivity and specificity typically above 95%, indicating robust predictive performance. Surprisingly, explainable AI approaches were infrequently applied, highlighting the persistent need for greater model transparency to foster clinical trust. Overall, these results demonstrate the strong potential of current AI based segmentation and classification pipelines. These methods can help clinicians accelerate the decision making process, increasing both the accuracy and efficiency of brain tumor diagnosis. These approaches may also support the development of personalized treatment plans tailored to each patient.

## 1. Introduction

Malignant brain tumors, although relatively rare, constitute one of the most lethal forms of cancer (DeAngelis, 2001). In 2022, Bray and colleagues (Bray et al., 2024) estimated the incidence and mortality of 36 cancer types across 185 countries, identifying brain tumors as the 19th in the incidence rank with 321,476 cases and the 12th in the mortality rank with 248,305 deaths. Brain tumors can be broadly categorized into primary or secondary types. The three most common types of primary brain tumors are meningioma, glioma, and pituitary tumors (Barnholtz-Sloan et al., 2018). Among these, meningiomas are the most frequent non-malignant tumors, while gliomas are the most common malignant ones, with glioblastoma being the most prevalent and aggressive subtype.

Gliomas originate from the glial cells, and they represent a heterogeneous group of tumors, ranging from low-grade gliomas (LGG) to very aggressive forms, staged as high-grade gliomas (HGG, i.e., glioblastoma) (Louis et al., 2021; Weller et al., 2024). LGGs grow slowly, while HGGs grow rapidly and are untreatable in some cases, even when advanced imaging, radiotherapy, and surgical techniques are employed. Gliomas may exhibit either well-circumscribed boundaries or diffusely infiltrative growth patterns, making segmentation and classification particularly challenging (Ohgaki and Kleihues, 2005). After tumor detection, an accurate diagnostic pipeline requires both tumor segmentation and classification before any therapeutic intervention.

To this end, among neuroimaging techniques, positron emission tomography (PET) can be employed to measure metabolic activity of brain tumors, and magnetic resonance imaging (MRI), provides critical information on the shape, size, location, and signal characteristics, essential for in-vivo diagnosis (Rasool and Bhat, 2025). MRI is considered the gold standard modality due to its non-invasive nature, its ability to provide high soft tissue contrast and spatial resolution (Heiss et al., 2011; Roy et al., 2013). Four standard MRI sequences are used for glioma diagnosis: T1-weighted MRI (T1), for discriminating healthy tissues and detect intralesional hemorrhages; T2-weighted MRI (T2), to delineate the edema region; T1-weighted MRI with gadolinium contrast enhancement (T1ce), to outline the more malignant part of the tumor; and Fluid-Attenuated Inversion Recovery (FLAIR), which helps distinguish edema from cerebrospinal fluid (Tjahyaningtijas, 2018). Therefore, a multiparametric MRI scan represents the most suitable approach to segment gliomas.

Glioma segmentation is crucial to remove confounding structures and provide accurate information for subsequent diagnosis. It allows for the anatomical delineation of brain tumors, differentiating neoplastic regions from normal brain areas that must be targeted by appropriate treatments. Additionally, segmentation enables monitoring of longitudinal MRI scans to track tumor recurrence, growth, or shrinkage (Dong et al., 2017; Işın et al., 2016). Glioma segmentation can be particularly challenging due to several factors, e.g. gliomas often exhibit isointense or hypointense characteristics on MRI, as well as poorly defined boundaries (Lather and Singh, 2020; Sachdeva et al., 2013). They also show significant inter-patient variability in terms of size, spatial extent, and anatomical location (Menze et al., 2015). Moreover, the presence of imaging artifacts and noise in MRI also increases the difficulty in segmentation (Huang et al., 2014). Several glioma subregions are clinically relevant and must be segmented: (i) core tumor (CT), defined as the combination of the non-enhancing tumor (NET), edema (ED), and necrotic regions (NCR) (Aumente-Maestro et al., 2025; Tejashwini et al., 2025), represents the central area containing the most aggressive cells, and is crucial for assessing tumor severity and guiding treatment and (ii) the enhancing tumor (ET), marked by active growth and increased vascularity, essential for understanding progression, planning surgery or radiotherapy, and estimating survival. Accurate segmentation of these regions is vital (Rasool and Bhat, 2025). Segmentation techniques can be classified, depending on the degree of human interaction required as defined by Olabarriaga et al. (2001), Yao (2005) and Foo (2006) (Foo, 2006; Olabarriaga and Smeulders, 2001; Yao, 2005), in manual, semi-automatic, and fully automatic. Manual segmentation is time-consuming, requires high expertise, is prone to error and operator-dependent (Gordillo et al., 2013). Semi-automatic methods require user input for initialization or correction of segmentation results (Gordillo et al., 2013). Fully automatic techniques require no human intervention. They are mainly based on model-based techniques that incorporate prior knowledge to solve the segmentation task, i.e., tumor shape, size, appearance, and location. This knowledge will be used as a starting point for the model fitting procedure, and will also allow expected variations in these characteristics (Gordillo et al., 2013).

Glioma classification is essential to ensure an accurate and reproducible identification of glioma subtypes (Louis et al., 2021). Histopathological classification is labor-intensive, requiring manual examination of both coarse and fine microscope image resolutions across large tissue volumes. When multiple pathologists classify the same sample, inconsistencies can arise due to subjective perception. Interobserver variability is a significant challenge, as prognostic outcomes can differ even within a single glioma subtype (Jin et al., 2021).

In this context, computational approaches such as artificial intelligence (AI) and radiomics, have emerged as complementary tools for glioma characterization. AI plays a key role by supporting and enhancing both glioma segmentation and classification, leveraging machine learning (ML) algorithms (Mitchell, 1999) to analyze radiomic features for tumor classification and deep learning (DL) models (Goodfellow et al., 2023) to automatically segment tumors and extract complex imaging patterns. Radiomics has emerged as a promising non-invasive approach. It extracts high-dimensional quantitative features from medical images, capturing tumor shape, texture, intensity, and heterogeneity patterns that may be imperceptible to the human eye. These features can accelerate and strengthen glioma classification, complementing histopathological assessment (Gillies et al., 2016; Lambin et al., 2012). DL is a subfield of ML that enables computational systems to learn from experience by constructing large neural network models with hierarchical representations of concepts capable of making accurate data-driven decisions (Goodfellow et al., 2023). In recent years, DL has rapidly become the most widely adopted computational approach in the field of ML, consistently outperforming traditional ML techniques across numerous domains, including medical image analysis. A major strength of DL lies in its ability to learn directly from large volumes of data, enabling models to achieve, and in some cases surpass, human-level performance on complex tasks (Alzubaidi et al., 2021). Most recent DL state-of-the-art approaches are based primarily on convolutional neural networks (CNNs). CNNs automatically learn and extract relevant features from raw imaging data, which is essential for complex tasks such as tumor segmentation and classification (Alzubaidi et al., 2021; Mlynarski et al., 2019). The most popular encoder-decoder CNN-based model in performing the segmentation tasks using medical imaging is U-Net (Ronneberger et al., 2015), which latent features can also be used for classification purposes.

This systematic review aims to critically evaluate and summarize the current state-of-the-art of DL-based approaches for glioma segmentation as the primary objective, and, where applicable, to further analyze studies that also address glioma classification as a subsequent step in the diagnostic pipeline. To this end, we established specific inclusion criteria to select relevant studies primarily focused on segmentation and, among these, we identified and examined those integrating a classification task. We then highlighted and compared their main characteristics, including the datasets used, imaging modalities, DL architectures, and segmentation and classification performance. Finally, we discuss the significance of achieving precise and accurate tumor segmentation, as a prerequisite for reliable classification.

## 2. Materials and Methods

### 2.1 Search and selection Criteria

This systematic review was conducted on published papers applying DL algorithms for the automatic segmentation of glioma through medical imaging. A study protocol outlining the search strategy and review methodology was preregistered in the Open Science Framework (OSF) database on the 2nd of December 2025 (Aresta, S., Palmirotta, C., Salvatore, C., & Asim, M. (2025, December 2). Advancing Brain Tumor Diagnosis Using Deep Learning: A Systematic Review on Glioma Segmentation and Classification on Multiparametric MRI. Retrieved from osf.io/8tvez). This work was performed and reported following Preferred Reporting Items for Systematic Reviews and Meta-Analyses (PRISMA) guidelines (Page et al., 2021). To identify the studies to be included in the review, the PICOS approach was used. Criteria for study inclusion and exclusion were established prior to the review. Papers were considered for inclusion only if (i) they were written in full-text English language in a peer-reviewed journal; (ii) they were published prior to our search on 14 March 2025; (iii) they were published from 01 January 2022 to 14 March 2025; (iv) they employed a DL algorithm for the segmentation task; (v) they specifically targeted the segmentation of glioma brain tumors; and (vi) they were fully accessible in full-text format. Studies were excluded if they met any of the following criteria: (a) they did not report segmentation performance metrics such as Dice Similarity Coefficient (DSC) or Intersection over Union (IoU); (b) they addressed only tumor detection or classification without segmentation; (c) they used AI-generated tumor masks as ground truth for training; (d) they were book chapters, review articles, surveys, or conference proceedings or (e) they were published before 01 January 2022. First, two authors independently screened the publications for the review (SA, CS). Papers that satisfied all predefined criteria were ultimately included in the systematic review. After identifying the includible papers, two authors (MA and SA) independently reviewed them to determine whether they performed a classification task. Studies were considered eligible for classification analysis only if they (i) used an AI algorithm to perform the classification task and (ii) reported classification performance metrics, such as accuracy. Any discrepancies between the reviewers were resolved through discussion and consensus.

### 2.2 Information Source and Search

A comprehensive search strategy was developed in collaboration with a librarian, incorporating both keywords and Medical Subject Headings (MeSH). One author (SA) carried out a literature search across the PubMed and Scopus databases. The search was conducted on 14 March 2025. The search strategy based on the PICOS method was applied following five concepts: (1) Population, defined as patients with glioma; (2) Intervention, defined as DL algorithms for glioma segmentation; (3) Comparison, defined as the manual segmentation of glioma; (4) Outcome, defined as the segmentation performance; (5) Study design, which should be “cross-sectional” or “longitudinal studies”. The search strategy was formed around four concepts: “brain tumor,” “glioma,” “segmentation,” and “deep learning”. Synonyms within each concept were combined with the OR Boolean operator, and terms between concepts were combined with the AND Boolean operator. The following keywords (with both extended names and abbreviations) were used for the literature search: ((“automatic segmentation” OR “segmentation”) AND (“deep learning” OR “DL”) AND (“brain” AND (“tumor” OR “tumour”)) AND (“detection”) AND (“glioma”)). No manual search of reference lists from previously retrieved articles was conducted by the authors.

### 2.3 Study Selection

Study selection was performed independently by two reviewers (SA, CP). After the deduplication procedure, initially the titles of the retrieved studies were screened, followed by abstract screening conducted by one reviewer (SA) assessing the abstracts for eligibility. The second reviewer (CP) independently revised the list of potential articles based on abstracts. Articles deemed potentially eligible were subsequently reviewed in detail by the same reviewers for quality assessment. Full-text evaluations were conducted to exclude studies that did not meet the inclusion criteria when eligibility could not be determined from the title and abstract alone. All studies reporting data suitable for pooling were included in the quantitative analysis. Specifically, we limited the analysis to studies that reported at least one performance metric of the automatic segmentation, including DSC or IoU.

From this pool of eligible studies, two reviewers (MA, SA) independently assessed whether the studies performed a classification task in addition to the segmentation task in full-text evaluations. Specifically, we included in the classification analysis only studies that reported at least one performance metric focused on tumor classification.

### 2.4 Data Extraction Strategy

The data extracted from each study were organized into the following categories: title, first author, year of publication, study objective, study population, tumor type, dataset used, sample size, imaging modality, image preprocessing techniques, use of external validation datasets, validation strategy, postprocessing techniques for segmentation masks, DL model employed for segmentation, model hyperparameters, segmented tumor regions, performance metrics used for model evaluation, use of explainable AI (XAI) methods, and reported performance results, i.e., study-specific DSC and study-specific IoU.

For studies that also performed the classification task, we additionally extracted: feature extraction method, feature selection, classification task performed, model employed for classification, hyperparameters for classification, classes considered, performance metrics used for model evaluation, any use of XAI method, and reported performance results, i.e., study-specific accuracy.

### 2.5. Risk of Bias in Individual Studies

In accordance with Cochrane guidelines, the Quality Assessment of Diagnostic Accuracy Studies (QUADAS) tool (Whiting et al., 2011) was employed to evaluate the methodological quality and risk of bias in each included study. Furthermore, the same tool was used for each paper also included in the classification pool.

Although QUADAS-2 was originally developed for diagnostic accuracy studies, it was adapted in this review to account for AI-based methodologies. In particular, patient selection was interpreted in terms of dataset provenance and sampling strategy; the index test domain was mapped to model development and validation procedures (including train–test separation and cross-validation); and the reference standard domain was evaluated based on annotation procedures and expert involvement. Flow and timing were assessed with respect to data splits and inclusion of all available samples in the final analysis.

Based on this assessment, studies were categorized as having low, high, or unclear risk of bias. Only studies classified as high-quality were included in the subgroup analyses for meta-analysis.

## 3. Results

### 3.1 Study Selection

All the stages of the selection process are shown in the PRISMA flow diagram in Figure 1. The literature search resulted in 310 papers from two electronic databases, no other papers were included in the list of previously retrieved papers. Before screening, 207 papers were excluded: 32 duplicate records; 32 reviews; 93 conference proceedings (i.e., conference papers and reviews); 5 book chapters, 2 surveys; 3 papers not written in full-text English (i.e., 1 written in Persian, and 2 written in Chinese), 2 papers not already published in peer-reviewed journals; and 37 papers published before 2022. A total of 104 papers were screened for title and abstract. 50 papers were excluded after the abstract and title screening. In particular, (i) 5 studies were excluded because they performed segmentation not using DL algorithms; (ii) 21 studies focused on detection of brain-tumor mass; (iii) 21 studies focused only on classification of brain tumors phenotypes; and (iv) 3 papers were out this review topic, one describing a quantitative system designed to standardise glioma imaging descriptions, another one mainly explaining theoretically EXplainable AI (XAI), and the latter focusing on the impact of two major programming environments on the accuracy of DL models for glioma detection. Therefore, at this screening phase, 53 papers were assessed in full-text for eligibility. At this step, 22 papers were excluded: (a) 4 papers were not available in full text, even after the authors’ request; (b) 11 papers did not provide DSC or IoU as segmentation performance metrics, but in the majority of the cases they computed only accuracy; (c) 6 papers performed segmentation not using DL methods, favoring the implementation of unsupervised clustering techniques, Wavelet filters, or region-growing algorithms; and (d) 1 paper was aimed at using annotation by clinicians and a Transformer-based model to generate the brain tumor segmentation masks to further train a Generative Adversarial Network (GAN) through a self-supervised approach. In the final stage, 31 studies met the eligibility criteria and were included in the review.

**Figure 1.**
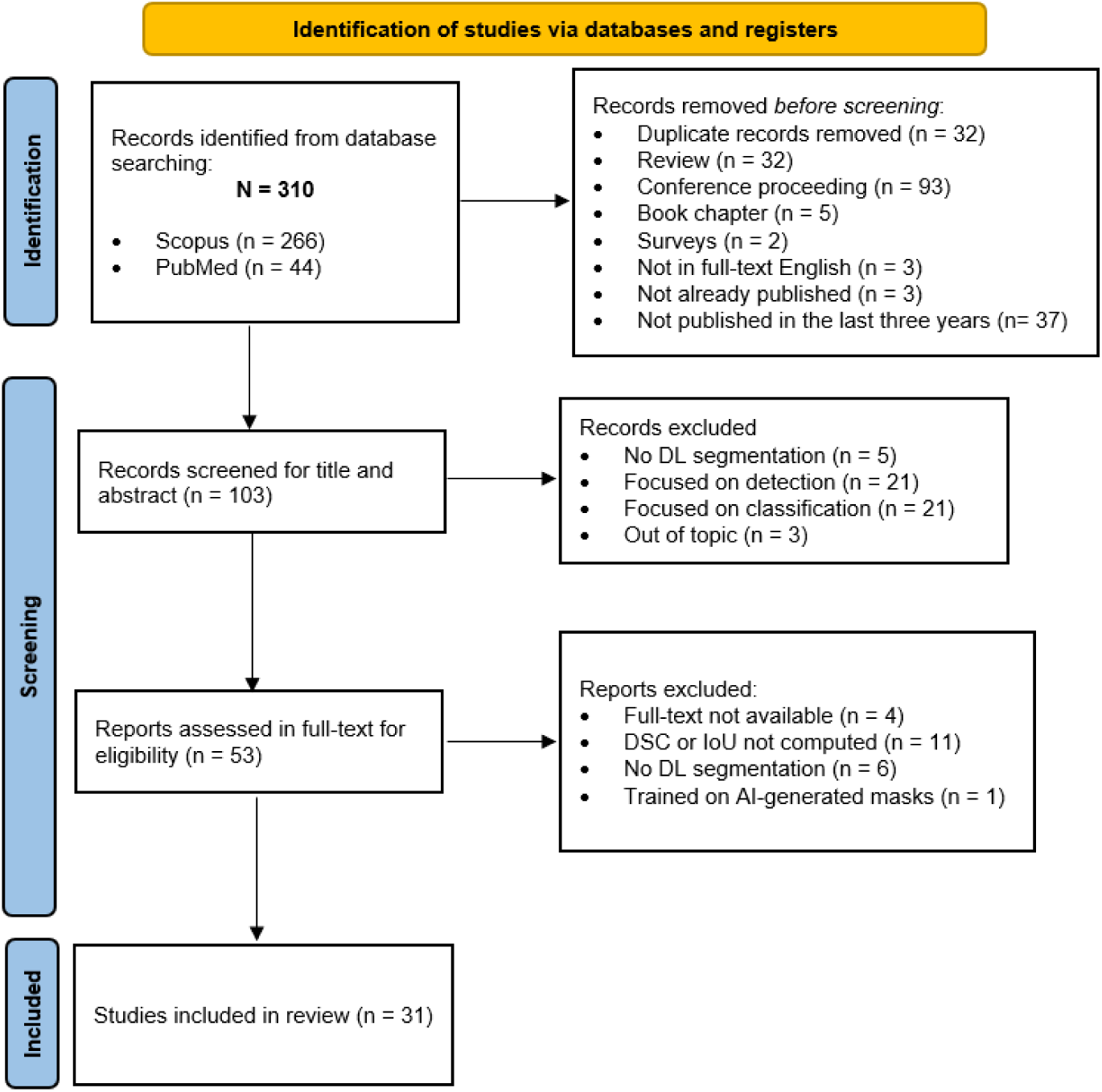
PRISMA flow chart outlining the sequential stages of the review selection process.

These papers were further screened in order to analyze how many papers not only performed segmentation using DL algorithms, but also performed a classification task using AI and provided performance metrics. Only 8 studies met this second eligibility criteria of classification and were further analyzed.

### 3.2 Results of the Systematic Review

#### 3.2.1 Brain Tumor Segmentation

Table S1 in the Supplementary Material summarizes the studies included in the systematic review and provides detailed information on the study population, tumor types under examination, dataset used, sample size, imaging modality for the segmentation task, image pre-processing techniques, segmentation mask post-processing, external validation or fine-tuning datasets, validation strategies for model training and testing, the DL model architecture, its hyperparameters, performance metrics used for evaluation (e.g., DSC, IoU, accuracy), the application of any XAI methods, and the main findings regarding model performance.

The final search yielded 31 studies. While most of these only focused on patients, three studies also included healthy controls (HC) (Krishnasamy and Ponnusamy, 2023; Mahajan et al., 2023; Shaikh et al., 2025). One study (Mahajan et al., 2023) focused specifically on the segmentation of pediatric brain tumors using DL algorithms. Three studies investigated segmentation for treatment-response assessment: two explored surgical treatment and one oral chemotherapy. Specifically, Bouget et al. (2022) (Bouget et al., 2022) trained a DL model using pre-operative MRI to segment brain tumors, aiming to improve the assessment of tumor size, location, and involvement of surrounding brain structures to optimize surgical planning. Zanier et al. (2023) (Zanier et al., 2023) developed a DL-based segmentation model trained on pre-operative MRI and fine-tuned on post-operative MRI to automatically compute tumor volume and estimate the extent of resection. In addition, Gutsche et al. (2023) (Gutsche et al., 2023) trained a DL algorithm on 18F-fluoroethyl-L-tyrosine PET (18F-FET PET) images from patients treated with adjuvant temozolomide following surgery in order to segment the Metabolic Tumor Volume (MTV) and predict disease prognosis and overall survival.

Among the selected studies, 21 focused exclusively on gliomas (Abd-Ellah et al., 2024; Aumente-Maestro et al., 2025; Bouget et al., 2022; Chetty et al., 2022; Esmaeilzadeh Asl et al., 2024; Koteswara Rao Chinnam et al., 2022; Kundal et al., 2024; Lefkovits et al., 2022; Mazher et al., 2024; Ottom et al., 2022; Rahat et al., 2023; Raza et al., 2023; Roy et al., 2023; Sachdeva et al., 2024; Tejashwini et al., 2025; Wang et al., 2022; Yang et al., 2025; Zafar et al., 2024; Zaitoon and Syed, 2023; Zaman et al., 2024; Zanier et al., 2023). Of these, four specifically addressed the segmentation of LGG only (Chetty et al., 2022; Ottom et al., 2022; Rahat et al., 2023; Zafar et al., 2024), while the remaining studies included both LGG and HGG. A smaller subset of studies (8 out of 31) also included other brain tumor types, such as meningiomas and pituitary tumors (Ahsan et al., 2025; Krishnasamy and Ponnusamy, 2023; Maqsood et al., 2022; Rai et al., 2024; Rosa et al., 2024; Shaikh et al., 2025; Sobhaninia et al., 2023; Xu et al., 2024). Furthermore, two studies (Gutsche et al., 2023; Mahajan et al., 2023), focused on a large variety of brain tumors and metastasis.

The majority of the papers used publicly available datasets, in particular, Brain Tumor Segmentation (BraTS) (Bakas et al., 2018, 2017; Menze et al., 2015), Brain Tumor Figshare (BTF) (Cheng, n.d.; Cheng et al., 2015), and The Cancer Genome Atlas – Lower Grade Glioma (TCGA-LGG) (Pedano et al., 2016). The BraTS dataset is a widely used, curated collection of multiparametric 3D brain MRIs from patients with both LGG and HGG. Each scan includes expert-approved manual segmentations of tumor subregions (whole tumor (WT), CT, ET), annotated by up to four raters following a standardized protocol and checked by experienced neuroradiologists. All images are preprocessed, i.e., co-registered, skull-stripped, resampled to 1 mm³ resolution, and intensity-normalized, for consistency and ease of use. A new BraTS dataset has been released every year from 2012 to the present, continuously expanding the number of subjects, improving data quality, and adding clinical information to support research on brain tumors. The BTF dataset includes T1ce-weighted MRIs of glioma, meningioma and pituitary tumors coupled with a binary mask indicating the WT generated as an average segmentation by 3 experienced raters. The TCGA-LGG dataset contains multiparametric MRI of LGG, coupled with WT segmentation generated using a semi-automatic method visually checked and modified if necessary by one experienced neuroradiologist. Delving into the included studies, 13 studies used only BraTS datasets from 2017 to 2021 (Abd-Ellah et al., 2024; Aumente-Maestro et al., 2025; Chetty et al., 2022; Esmaeilzadeh Asl et al., 2024; Koteswara Rao Chinnam et al., 2022; Lefkovits et al., 2022; Mazher et al., 2024; Raza et al., 2023; Roy et al., 2023; Tejashwini et al., 2025; Wang et al., 2022; Zaitoon and Syed, 2023; Zaman et al., 2024), 4 studies used only BTF dataset (Rai et al., 2024; Rosa et al., 2024; Sobhaninia et al., 2023; Xu et al., 2024), and 3 studies used only TCGA-LGG (Ottom et al., 2022; Rahat et al., 2023; Zafar et al., 2024). Furthermore, 3 papers used a combination of these datasets (Ahsan et al., 2025; Maqsood et al., 2022; Yang et al., 2025). Other less prominent datasets from Kaggle were also used in a small subset, such as the Kaggle Brain Tumor MRI dataset (Nickparvar, 2021) and the Kaggle Brain Classification dataset (Bhuvaji et al., 2020), either independently (Krishnasamy and Ponnusamy, 2023) or in combination with BraTS datasets (Shaikh et al., 2025). In particular, one study (Kundal et al., 2024) used BraTS datasets in combination with UPENN-GBM dataset (Bakas et al., 2021). Additionally, 5 studies used custom-made datasets independently (Gutsche et al., 2023; Mahajan et al., 2023) or in combination with BraTS datasets (Bouget et al., 2022; Sachdeva et al., 2024; Zanier et al., 2023).

With regard to population size, studies relied either on a single dataset or on a combination of multiple datasets. In the latter case, the total number of patients was computed as the sum of the cohorts from each dataset. Based on this approach, the median number of patients was 369, ranging from 65 to 3,264.

Looking at the image modalities, only one paper used 18F-FET PET images (25), the other 30 papers used MRI. Specifically, 7 studies used T1ce-weighted MRI (Ahsan et al., 2025; Krishnasamy and Ponnusamy, 2023; Maqsood et al., 2022; Rai et al., 2024; Rosa et al., 2024; Sobhaninia et al., 2023; Xu et al., 2024), 3 studies used FLAIR MRI images (Ottom et al., 2022; Rahat et al., 2023; Zafar et al., 2024), 13 studies used FLAIR, T1-weighted, T1ce-weighted and T2-weighted MRI (Abd-Ellah et al., 2024; Aumente-Maestro et al., 2025; Chetty et al., 2022; Esmaeilzadeh Asl et al., 2024; Kundal et al., 2024; Lefkovits et al., 2022; Mazher et al., 2024; Raza et al., 2023; Roy et al., 2023; Shaikh et al., 2025; Tejashwini et al., 2025; Yang et al., 2025; Zaitoon and Syed, 2023), 1 used FLAIR, T1-weighted, T1ce-weighted and T2-weighted and Diffusive-weighted imaging (DWI) MRI (Mahajan et al., 2023), 1 used FLAIR and T2-weighted MRI (Koteswara Rao Chinnam et al., 2022), 2 used FLAIR and T1ce-weighted MRI (Bouget et al., 2022; Zanier et al., 2023), 2 used FLAIR, T1ce-weighted and T2-weighted MRI (Sachdeva et al., 2024; Zaman et al., 2024). Only one paper used MRI without reporting any other information (Wang et al., 2022).

Image pre-processing techniques are essential when dealing with DL image segmentation, with considerable variability in techniques depending on the dataset employed. The included studies applied resizing to ensure consistent image dimensions (Abd-Ellah et al., 2024; Ahsan et al., 2025; Bouget et al., 2022; Chetty et al., 2022; Esmaeilzadeh Asl et al., 2024; Koteswara Rao Chinnam et al., 2022; Krishnasamy and Ponnusamy, 2023; Ottom et al., 2022; Rahat et al., 2023; Rai et al., 2024; Raza et al., 2023; Rosa et al., 2024; Sachdeva et al., 2024; Shaikh et al., 2025; Wang et al., 2022; Xu et al., 2024; Zaitoon and Syed, 2023; Zaman et al., 2024), intensity normalization to facilitate model training (Abd-Ellah et al., 2024; Aumente-Maestro et al., 2025; Bouget et al., 2022; Chetty et al., 2022; Gutsche et al., 2023; Koteswara Rao Chinnam et al., 2022; Krishnasamy and Ponnusamy, 2023; Mahajan et al., 2023; Rahat et al., 2023; Rai et al., 2024; Raza et al., 2023; Sachdeva et al., 2024; Wang et al., 2022; Zaitoon and Syed, 2023; Zanier et al., 2023), data augmentation to expand the dataset and enhance generalization (Abd-Ellah et al., 2024; Aumente-Maestro et al., 2025; Bouget et al., 2022; Gutsche et al., 2023; Koteswara Rao Chinnam et al., 2022; Krishnasamy and Ponnusamy, 2023; Mahajan et al., 2023; Rahat et al., 2023; Rai et al., 2024; Roy et al., 2023; Shaikh et al., 2025; Zanier et al., 2023), image cropping to manage computational load and focus on local image features (Abd-Ellah et al., 2024; Aumente-Maestro et al., 2025; Chetty et al., 2022; Gutsche et al., 2023; Koteswara Rao Chinnam et al., 2022; Krishnasamy and Ponnusamy, 2023; Sobhaninia et al., 2023), and noise filtering to reduce acquisition artifacts and improve the distinction of anatomical structures (Esmaeilzadeh Asl et al., 2024; Krishnasamy and Ponnusamy, 2023; Rosa et al., 2024; Sobhaninia et al., 2023; Zaitoon and Syed, 2023; Zaman et al., 2024). Furthermore, 3 studies applied post-processing techniques to DL segmented masks, applying morphological operations (Maqsood et al., 2022), resampling and resize (Mazher et al., 2024), thresholding and connected component labeling technique (CCL) (Rai et al., 2024).

To train and test the DL model, 22 papers used the hold-out validation approach with different percentage of splitting (Abd-Ellah et al., 2024; Chetty et al., 2022; Esmaeilzadeh Asl et al., 2024; Koteswara Rao Chinnam et al., 2022; Krishnasamy and Ponnusamy, 2023; Lefkovits et al., 2022; Mahajan et al., 2023; Mazher et al., 2024; Ottom et al., 2022; Rahat et al., 2023; Rai et al., 2024; Raza et al., 2023; Rosa et al., 2024; Roy et al., 2023; Sachdeva et al., 2024; Shaikh et al., 2025; Tejashwini et al., 2025; Xu et al., 2024; Yang et al., 2025; Zafar et al., 2024; Zaitoon and Syed, 2023; Zaman et al., 2024), 3 studies used the cross-validation approach (Bouget et al., 2022; Sobhaninia et al., 2023; Zanier et al., 2023), and 4 papers combined the two aforementioned validation approaches (Ahsan et al., 2025; Aumente-Maestro et al., 2025; Gutsche et al., 2023; Wang et al., 2022). Only 1 study used pre-trained models (Kundal et al., 2024), and 1 study did not provide any information (Maqsood et al., 2022). Furthermore, 9 studies performed external validation (Ahsan et al., 2025; Gutsche et al., 2023; Koteswara Rao Chinnam et al., 2022; Kundal et al., 2024; Raza et al., 2023; Shaikh et al., 2025; Yang et al., 2025; Zafar et al., 2024; Zanier et al., 2023).

Regarding the DL model hyperparameters, the most commonly used loss function is the Dice loss, 3 studies combined with Binary Cross-Entropy loss to improve segmentation stability and accuracy (Rai et al., 2024; Shaikh et al., 2025; Tejashwini et al., 2025). Moreover the most used activation functions were the Rectified Linear Unit (ReLU) and sigmoid functions. The most used optimizers were the Adaptive Moment Estimation (Adam) and the Stochastic Gradient Descent (SGD), in particular 4 out 31 studies used SGD optimizer (Abd-Ellah et al., 2024; Ahsan et al., 2025; Lefkovits et al., 2022; Sobhaninia et al., 2023), 15 out of 31 used Adam optimizer (Bouget et al., 2022; Chetty et al., 2022; Esmaeilzadeh Asl et al., 2024; Krishnasamy and Ponnusamy, 2023; Maqsood et al., 2022; Ottom et al., 2022; Rai et al., 2024; Raza et al., 2023; Rosa et al., 2024; Sachdeva et al., 2024; Shaikh et al., 2025; Tejashwini et al., 2025; Wang et al., 2022; Zaitoon and Syed, 2023; Zaman et al., 2024), and 2 studies used both (Koteswara Rao Chinnam et al., 2022; Roy et al., 2023). The learning rate used ranged from 1E-08 (Wang et al., 2022) to 0.9 (Roy et al., 2023), and the batch size ranged from 1 to 128 (Maqsood et al., 2022). Only 3 papers did not provide the hyperparameters used (Gutsche et al., 2023; Kundal et al., 2024; Rahat et al., 2023).

DL models were trained to segment one or several tumor regions. Furthermore, 1 study was trained to segment metabolic tumor volume (MTV) (Gutsche et al., 2023), 13 studies segmented only the WT volume (Ahsan et al., 2025; Bouget et al., 2022; Krishnasamy and Ponnusamy, 2023; Maqsood et al., 2022; Ottom et al., 2022; Rahat et al., 2023; Rai et al., 2024; Rosa et al., 2024; Roy et al., 2023; Sobhaninia et al., 2023; Xu et al., 2024; Zafar et al., 2024; Zaman et al., 2024), 9 studies segmented the WT, CT, and ET (Abd-Ellah et al., 2024; Aumente-Maestro et al., 2025; Chetty et al., 2022; Koteswara Rao Chinnam et al., 2022; Mazher et al., 2024; Raza et al., 2023; Sachdeva et al., 2024; Tejashwini et al., 2025; Yang et al., 2025), 2 studies segmented WT, ET, and NET (Mahajan et al., 2023; Zanier et al., 2023), one study segmented NCR, ET and ED (Esmaeilzadeh Asl et al., 2024), and another one segmented these plus NET (Zaitoon and Syed, 2023). In addition, 1 paper segmented ET, NET and ED (Kundal et al., 2024), 1 study segmented only the WT using specific datasets, while WT, CT, and ET were segmented using other datasets (Shaikh et al., 2025), and the remaining one segmented healthy tissue, WT, ED, active tumor, and NCR/NET regions (Lefkovits et al., 2022). Lastly, only 1 study did not report the segmented regions (Wang et al., 2022)

The specific approaches and performances of the individual studies are summarized below, highlighting the diversity of DL architectures and segmentation strategies applied across datasets and tumor subregions. Wang et al. (2022) (Wang et al., 2022) proposed a multiparametric DL framework named Encoder–Decoder with semantic gap compensation Unit (E-DU) for brain tumor segmentation. The model is based on an encoder–decoder CNN architecture designed to address the semantic gap between low-level and high-level features, Specifically, E-DU incorporates a semantic gap compensation module that refines and aligns feature extracted at different network depths, improving the consistency between encoder and decoder pathways. The proposed model achieved a DSC of 95.31%.

To assess treatment response, Gutsche et al. (2023) (Gutsche et al., 2023) developed a DL–based method, based on nnU-Net (Isensee et al., 2021) to segment MTV. This self-configuring architecture automatically adapts preprocessing, network design, and training parameters to the dataset, ensuring data-drive performance optimization without manual tuning. The model achieved DSC values of 80.00% and 81.00%, with and without prior brain extraction, respectively. Performance was further evaluated across lesion sizes, yielding DSCs of 66.50%, 90.00%, and 87.00% for small, medium, and large lesions, respectively. Finally, nnU-Net was applied to segment single, non-malignant, and multifocal lesions, achieving DSCs of 83.00%, 77.00%, and 67.00%, respectively.

Focusing on those papers that segmented only the WT, Ahsan et al. (2024) (Ahsan et al., 2025) used a combination of YOLOv5 for detection with a 2D U-Net for tumor segmentation. The model achieved a DSC of 88.10% for segmentation, outperforming both standalone 2D U-Net (DSC=80.50%) and Mask R-CNN (DSC=44.2%). Maqsood et al. (2022) (Maqsood et al., 2022) developed a 17-layer CNN to perform the automatic segmentation task. The CNN was designed to hierarchically extract relevant features of tumor tissue, generating precise segmentation maps prior to the classification stage. This model achieved a DSC of 96.71% on BraTS2018 and of 97.87% on BTF. Furthermore, it was the only study that applied Gradient-weighted Class Activation Mapping (Grad-CAM) for segmentation model explanation. The Grad-CAM analysis confirmed that the network focused on the tumor boundaries and heterogeneous internal regions, highlighting its ability to identify clinically relevant features. With the same aim of improving focus on local regions in a pre-post operative setting, Bouget et al. (2022) (Bouget et al., 2022) developed the Attention-Gate U-Net (AGU-Net). This model extends the standard U-Net by incorporating attention gates, modules designed to highlight the most relevant regions in the input image while suppressing irrelevant background. The model demonstrated strong performance, achieving a DSC of 86.00% for HGG and 75.00% for LGG.

Following a similar approach, Rai et al. (2024) (Rai et al., 2024) employed a modified U-Net architecture with an EfficientNet encoder backbone (evaluating variants from B0 to B5) ensuring precise localization and delineation of tumor regions. The network comprises two heads: one for segmentation, and one for classification. Both heads share the EfficientNet encoder, branching only at task-specific layers, allowing the model to leverage shared features for both tasks. The segmentation performance varied across EfficientNet variants. Without post-processing, the highest DSC were achieved by B4 and B5 (i.e., 83.50%), followed by B1 (82.56%), B3 (82.07%), B0 (80.79%), and B2 (74.22%). With post-processing, B4 again performed best (87.00%), followed by B1 (86.62%), B5 (85.95%), B3 (84.20%), B0 (83.35%), and B2 (77.19%). In the same way, Sobhaninia et al. (2023) proposed a Multiscale Cascaded Multitask Network based on a U-Net backbone with multiscale feature aggregation, designed to perform tumor segmentation and classification simultaneously. The network processes multiple resolution levels in a cascaded manner, capturing both fine and coarse features, while a feature aggregation module fuses information from different levels. The Multiple Cascaded Multitask Network reached a DSC of 95.93%, and adding feature aggregation further improved it to 96.27%, demonstrating the effectiveness of combining multiscale, cascaded, and aggregation strategies for precise tumor segmentation. Also Roy et al. (2023) (Roy et al., 2023) proposed two lightweight U-Net–based architectures for brain tumor segmentation, named S-Net and SA-Net. Both models retain the classic U-Net, but they reduce computational overhead by using residual blocks with a single convolution per level and by implementing a simplified merge strategy. SA-Net further incorporates an attention block after merging, which emphasizes salient tumor features while suppressing irrelevant regions. For S-Net, the best performance was achieved on LGG after 100 epochs, with a DSC of 81.03%, while for HGG the peak DSC was 77.56%. For SA-Net, the highest DSC was observed on LGG after 100 epochs, with a DSC of 81.20%, and for HGG, DSC reached 77.80%. In addition, Zafar et al. (2024) (Zafar et al., 2024) introduced Enhanced TumorNet, a hybrid DL model combining YOLOv8s and U-Net for brain tumor detection and segmentation, respectively. U-Net reached an IoU of 80.00%. Furthermore, Rosa et al. (2024) (Rosa et al., 2024) investigated how different image denoising techniques, such as median, Gaussian, anisotropic diffusion, and bilateral filtering, affect the performance of U-Net. Bilateral filtering provided the best balance between computational efficiency and segmentation accuracy, achieving a DSC of 76.96%.

Not all the included studies proposed a U-Net based model to segment the WT. Xu et al. (2024) (Xu et al., 2024) implemented a different approach called Deep-GARL, a hybrid model combining Generative Adversarial Networks (GANs) and Reinforcement Learning (RL) to improve segmentation of low-contrast and small brain structures. Using a semi-supervised approach, the model leverages both labeled and unlabeled data, while the RL component directs attention to relevant regions. This model reached a DSC of 85.02%. Krishnasamy and Ponnusamy (2023) (Krishnasamy and Ponnusamy, 2023) proposed two hybrid DL models: Model 1 used a Fully Convolutional Networks (FCN)-32 architecture for segmentation, followed by a ResNet-50 classifier, while Model 2 employed SegNet for segmentation with MobileNet as classifier. The FCN-32 (Long et al., 2014) architecture upsamples feature maps to the original image resolution, enabling accurate delineation of tumor regions whereas SegNet (Badrinarayanan et al., 2015) employs an encoder–decoder structure with pooling indices to efficiently reconstruct high-resolution segmentation maps from low-resolution feature representations. FCN-32 slightly outperformed SegNet, with a DSC, respectively equal to 77.40% and 76.00%. Rahat et al. (2023) (Rahat et al., 2023) integrated several DL models, i.e., DeepLabv3, U-Net, DenseNet121-Unet, ResNet50, Attention U-Net, and EfficientNet, to enhance segmentation accuracy. Among these, EfficientNet achieved the highest performance with a DSC of 92.37%, followed by DenseNet121 (DSC=85.23%) and DeepLabv1 (DSC=84.13%).

Focusing on WT, CT, ET segmentations, Chetty et al. (2022) (Chetty et al., 2022) used a 3D U-Net reaching a DSC for, respectively, WT, CT and ET equal to 94.00%, 90.00% and 84.00%. Koteswara Rao Chinnam et al. (2022) (Koteswara Rao Chinnam et al., 2022) introduced two closely-related architectures: Multimodal Cascaded Convolution U-Net (MCC-Net) and Multimodal Attention Cascaded U-Net (MAC-Net). MCC-Net integrates attention gates, skip connections, and group normalization to capture salient features across multiple MRI modalities, providing stability and improved performance. MAC-Net is the same architecture without group normalization, serving as a baseline to assess the effect of normalization. Evaluated on the BraTS 2018 dataset, MCC-Net achieved DSCs of 94.47% for WT, 84.12% for CT, and 82.72% for ET in HGG, while for LGG it obtained DSCs of 85.71% for WT, 78.85% for CT, and 74.16% for ET.

Later, Mazher et al. (2023) (Mazher et al., 2024) introduced Residual Network with Multi-Head Attention (ResMHA-Net) to capture complex spatial and contextual features. ResMHA-Net achieved comparable DSC, equal to 90.20% for the WT, 89.10% for CT, and 88.30% for the ET. In the same year, Raza et al. 2023 using a Residual U-Net (dResU-Net), which aimed to preserve low-level features while capturing high-level contextual information, achieved strong segmentation performance across multiple BraTS datasets. On BraTS 2020, dResU-Net reached DSCs of 92.12% for WT, 86.60% for CT, and 80.04% for ET. Similarly, on BraTS 2021, it obtained DSCs of 86.01% for WT, 84.00% for CT, and 82.21% for ET.

Furthermore, Abd-Ellah et al. (2024) (Abd-Ellah et al., 2024) implemented Two Parallel Cascaded U-Nets with Asymmetric Residual-blocks Network (TPCUAR-Net) to effectively capture multi-scale features by combining symmetric U-Net architecture with asymmetric residual-blocks. The model leverages skip connections and residual units to enhance feature learning and boundary delineation, obtaining a DSC, respectively, for WT, CT, and ET, equal to 91.00%, 83.00% and 80.00%. Zaman et al. (2024) (Zaman et al., 2024) using Adaptive Feature Medical Segmentation Network (AFMS-Net). The network can incorporate two different encoder modules: the Single Adaptive Encoder Block (SAEB) for efficiency and the Dual Adaptive Encoder Block (DAEB) for complex feature extraction, along with advanced attention mechanisms to capture spatial and channel-wise dependencies. For AFMS-SAEB, the model achieved a DSC equal to 89.40%. For individual regions, DSCs were 88.00% for WT, 85.00% for CT, and 83.00% for ET. AFMS-DAEB slightly improved these results, reaching an overall DSC of 90.20%, with region-based DSCs of 87.00% for WT, 87.00% for CT, and 85.0% for ET.

Recently, Tejaswini et al. (2025) (Tejashwini et al., 2025) improved the standard U-Net framework for automatic brain tumor segmentation by incorporating residual dense blocks, layered attention mechanisms, and class-specific attention modules. This model, called Scleral Residue Class Attention U-Net (SLCA-UNet), achieved excellent segmentation performance across multiple BraTS datasets. On BraTS 2017, it obtained a WT DSC of 91.50%, CT DSC of 85.20%, and ET DSC of 93.10%. Similar results were reported for BraTS 2018 (WT DSC=91.50%, CT DSC=85.20%, ET DSC=93.10%), BraTS 2019, and BraTS 2020. Also Aumente-Maestro C. et al. (2025) (Aumente-Maestro et al., 2025) introduced Brain Tumor Segmentation (BTS) U-Net, a lightweight DL architecture based on U-Net. This model enhances the standard U-Net framework by incorporating residual dense blocks and a class-attention mechanism. For HGG using 10-fold cross-validation, it achieved the highest DSC values of 91.10% for the WT, compared to the other subregions, i.e., 89.50% for the CT, and 84.80% for the ET. When trained exclusively on HGG cases, BTS U-Net further improved its performance, reaching DSCs of 91.20% for WT, 91.00% for CT, and 85.40% for ET. Yang et al. (2025) (Yang et al., 2025) introduced MUNet, which combines UNet and Mamba networks (Gu and Dao, 2023) for accurate brain tumor segmentation. It incorporates Selective Scanning-State Space Model (SD-SSM) and Spatial and Channel Reconstruction Convolution (SD-Conv) modules to enhance feature extraction and computational efficiency. Evaluated on BraTS datasets, MUNet achieved DSCs of 81.50% (ET), 90.10% (WT), 81.50% (CT) on BraTS 2018, 83.50% (ET), 91.50% (WT), 82.30% (CT) on BraTS 2020, and 70.20% DSC on TCGA-LGG. Lastly, Shaikh et al. (2025) (Shaikh et al., 2025) investigated multiple DL models for glioma segmentation across several datasets. Compared to the other dataset used to train the model, the pretrained Visual Geometry Group (VGG)-16 (Simonyan and Zisserman, 2014) achieved the highest DSC of 98.03% when using the Kaggle Brain Tumor MRI dataset. For the BraTS 2020 dataset, the pretrained 3D Squeeze-and-Excitation DenseNet201 (SEL-DenseNet201) obtained a DSC of 94.87% for the WT. In BraTS 2021, SEL-DenseNet201 again performed best with DSCs of 96.00% for ET, 93.02% for CT, and 93.66% for WT. Finally, on BraTS 2023, SEL-DenseNet201 reached the highest DSC values of 96.10% for ET, 93.21% for TC, and 93.78% for WT compared to the other employed datasets.

Regarding the segmentation of WT, ET, and NET, Mahajan A. et al. (2023) (Mahajan et al., 2023) employed a variation of 3D U-Net to segment several pediatric tumors, achieving DSCs of 61.00% for WT, 59.00% for ET, and 31.00% for NET. Improved performance in a treatment setting was reported by Zaneir et al. (2023) (Zanier et al., 2023), who used a standard 2D U-Net in both pre-and post-operative segmentation settings. In the pre-operative scenario, the model achieved DSCs of 73.00% for WT, 62.00% for ET, and 43.00% for NET. In the post-operative scenario, performance decreased, with DSCs of 59.00% for WT, 21.00% for ET, and 7.00% for NET.

Esmaeilzadeh Asl et al. (2024) (Esmaeilzadeh Asl et al., 2024) proposed a method using hybrid filters to enhance the feature extraction capabilities of the U-Net model for segmenting NCR, ED, and ET regions, achieving an overall DSC of 87.39%. Building on U-Net, Zaitoon and Sayed (2023) (Zaitoon and Syed, 2023) employed 3D Residual Recurrent U-Net 2+ (3D RR-UNet2+), a 3D residual recurrent U-Net designed to capture both spatial and contextual information, to segment the same tumor subregions, while also including NET. This model demonstrated high segmentation accuracy in HGG cases, with DSCs of 98.39% for NCR, 99.10% for ED, 98.69% for ET, and 98.20% for NET. In LGG cases, performance remained excellent, with DSCs of 98.32% for NCR, 98.94% for ED, 98.94% for ET, and 99.41% for NET. In parallel, a comparative study of Kundal et al. (202) (Kundal et al., 2024) evaluated the segmentation of ET, NET, and ED using 3D CaPTk, 2DVNet, 3D EnsembleUNets, and ResNet50. CaPTk is a 3D CNN-based platform for quantitative image analysis, 2DVNet is a 2D volumetric U-Net variant, EnsembleUNets combines multiple U-Net architectures for robust segmentation, and ResNet50 is a 2D residual CNN adapted for medical imaging. On the BraTS 2021 dataset, 3D CaPTk achieved a DSC of 91.00%, 2DVNet and ResNet50 performed poorly with DSCs of 0.60%, and EnsembleUNets achieved the highest DSC of 93.00%. On the UPENN-GBM dataset, CaPTk reached a DSC of 84.00%, EnsembleUNets 85.00%, while 2DVNet and ResNet50 again showed minimal performance (1.00% DSC), highlighting the superior accuracy and robustness of 3D CaPTk and EnsembleUNets for brain tumor segmentation.

Finally, Lefkovits et al. (2022) (Lefkovits et al., 2022) employed a 3D encoder-decoder architecture for glioma segmentation, utilizing various pretrained models. Specifically, they integrated ResNet50 and ResNet101 as encoders with decoder architectures such as FCN, Pyramid Scene Parsing Network (PSPNet), and DeepLabv3+. Additionally, they implemented stacking ensemble learning (SEL) to enhance model performance. In their experiments on the Kaggle Brain Tumor MRI dataset, the best single model was VGG-16 achieving a DSC of 98.03%, while the SEL-DenseNet201 model achieved a DSC of 97.43%

To provide a clearer overview of DSC values, Figure 2 illustrates the distributions of overall DSC scores as well as those across different tumor regions, including WT, CT, and ET. The median overall DSC was 80.73%, ranging from 0.60% to 98.59%. Considering glioma subregions, the median DSC for WT was 89.20% (range: 59.00–94.87%), for CT it was 83.56% (range: 64.26–92.30%), and for ET it was 80.04% (range: 21.00–98.94%).

**Figure 2.**
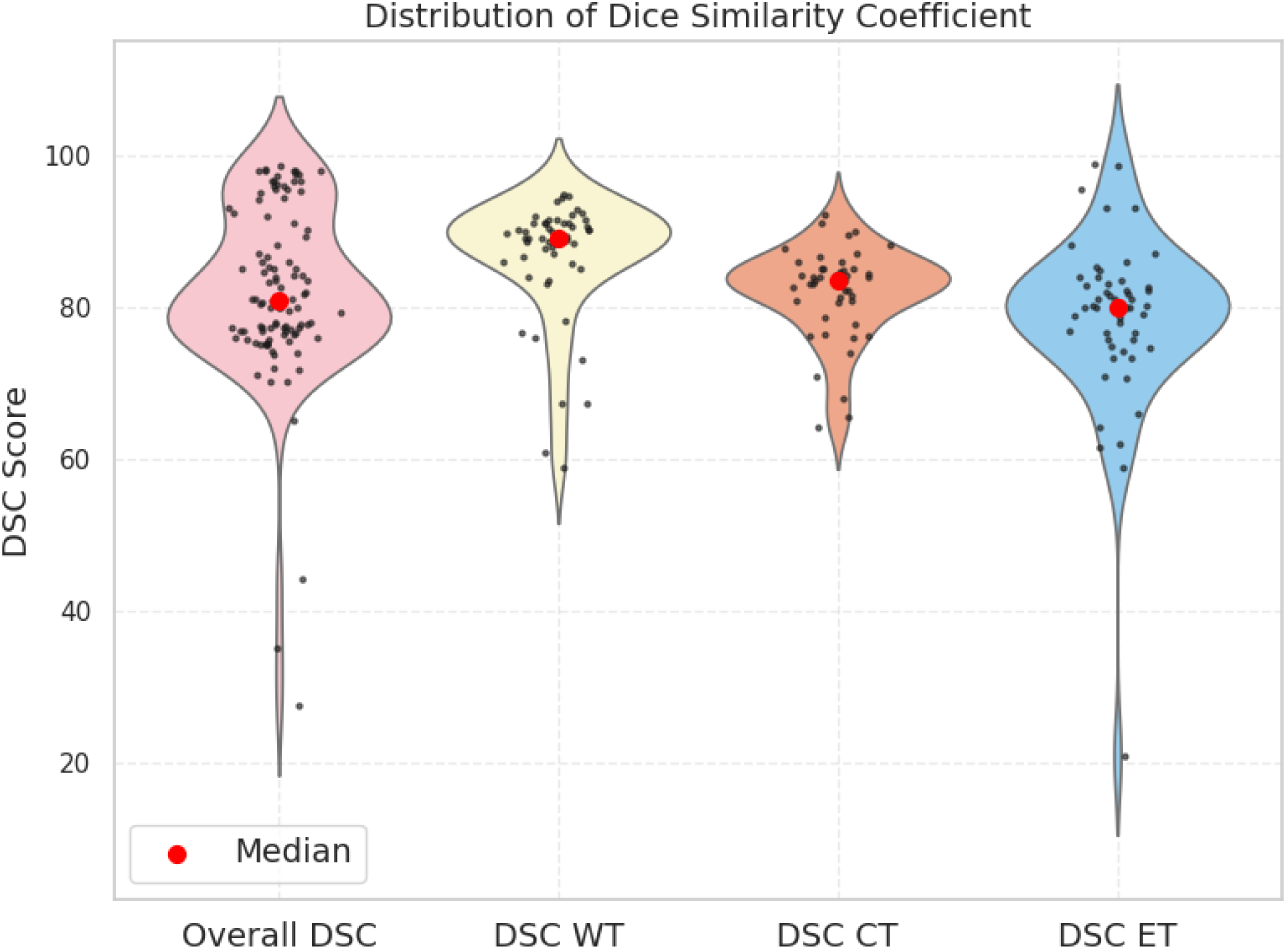
Violin plots were used to visually display the distributions of the overall DSC scores but also across different tumor regions, including WT, CT, and ET. Each violin represents the distribution of all available DSC values reported. Black dots indicate individual model performances, while the red dots highlight the median of each distribution. This representation allows for a clear comparison of performance variability and central tendency across tumor regions, capturing both the spread and the typical values of the reported results. *Legend: DSC, Dice Score Coefficient; WT, whole tumor; CT, core tumore; ET, enhancing tumor*

#### 3.2.2 Brain Tumor Classification

Table S2 in the Supplementary Material summarizes the studies that performed segmentation followed by classification, thus providing additional information related to the classification task. In particular, the feature extraction method and the features used for the classification, the feature selection algorithm applied (if any), the training/validation/testing splits, the presence of external validation or fine-tuning datasets, the model hyperparameters, the target of the classification task (i.e., the predicted classes), the performance metrics used for classification, the application of XAI methods, and the major findings obtained.

Among the 31 identified studies eligible for the systematic review, 8 focused on both brain-tumor segmentation and classification (Abd-Ellah et al., 2024; Aumente-Maestro et al., 2025; Krishnasamy and Ponnusamy, 2023; Mahajan et al., 2023; Maqsood et al., 2022; Rai et al., 2024; Sobhaninia et al., 2023; Zaitoon and Syed, 2023), while the remaining studies addressed only the segmentation task. Among these studies, two included both patients and controls (Krishna et al., 2025; Mahajan et al., 2023). Regarding the tumor type, 3 focused their attention on glioma (Abd-Ellah et al., 2024; Aumente-Maestro et al., 2025; Zaitoon and Syed, 2023), in particular on HGG and LGG, 4 studies investigated glioma, meningioma and pituitary tumors (Krishna et al., 2025; Maqsood et al., 2022; Rai et al., 2024; Sobhaninia et al., 2023), and only 1 study considered diverse tumor cases, including brainstem glioma (Mahajan et al., 2023).

To perform segmentation and classification tasks, 2 studies used only the BTF dataset (Rai et al., 2024; Sobhaninia et al., 2023), 3 used BraTS from 2017 to 2020 (Abd-Ellah et al., 2024; Aumente-Maestro et al., 2025; Zaitoon and Syed, 2023), one used both BTF and BraTS 2018 (Maqsood et al., 2022), another one used the Kaggle Brain Tumor MRI Dataset (Krishnasamy and Ponnusamy, 2023), and the last one used a custom-made not publically-available dataset (Mahajan et al., 2023).

As previously demonstrated, studies used a single dataset or a combination of datasets, reaching a median sample size of 301, with a minimum number of subjects included equal to 94, and a maximum number of subjects equal to 3264.

All selected papers used MRI, though the specific modalities varied across studies. Three studies employed T1ce-weighted MRI (Maqsood et al., 2022; Rai et al., 2024; Sobhaninia et al., 2023), other 3 studies used multiparametric MRI, i.e., FLAIR, T1-weighted, T1ce-weighted and T2-weighted (Abd-Ellah et al., 2024; Aumente-Maestro et al., 2025; Zaitoon and Syed, 2023), and 1 study also added DWI sequence (Mahajan et al., 2023). Only 1 study did not specify which sequences were used (Krishnasamy and Ponnusamy, 2023).

Features used to train the classification model, in 7 out of 8 papers, were extracted using a CNN model (Abd-Ellah et al., 2024; Krishna et al., 2025; Mahajan et al., 2023; Maqsood et al., 2022; Rai et al., 2024; Sobhaninia et al., 2023; Zaitoon and Syed, 2023). Maqsood S. et al. (2022) (Maqsood et al., 2022) used MobileNetV2 (Sandler et al., 2018) a CNN model with inverted residuals and linear bottlenecks, to extract low-, middle-, and high-level features for classification; the extracted features were subsequently refined through feature selection using an entropy-controlled method. The other six studies employed a single CNN-based DL model to perform both segmentation and classification, leveraging the same feature representations extracted by the CNN for training and prediction in both tasks (Abd-Ellah et al., 2024; Krishna et al., 2025; Mahajan et al., 2023; Rai et al., 2024; Sobhaninia et al., 2023; Zaitoon and Syed, 2023). Among these, 1 study combined CNN features and clinical information only to perform classification (Mahajan et al., 2023). Furthermore, Aumente-Maestro et al. (2025) (Aumente-Maestro et al., 2025), after the segmentation extracted radiomics features, that were clustered in MRI features i.e., mean, standard deviation, minimum and maximum pixel intensity, skewness, kurtosis, entropy, and blurriness, and features mainly related to the segmented mask called segmented, i.e., the number of pixels assigned to each segmented area, the tumor size and extension along each of the X, Y, and Z axes.

The validation strategies reflected different levels of rigor in model evaluation. Specifically, 5 studies used a hold-out validation approach, using different percentages for train, validation, and test sets splitting (Abd-Ellah et al., 2024; Krishnasamy and Ponnusamy, 2023; Mahajan et al., 2023; Rai et al., 2024; Zaitoon and Syed, 2023). More rigorous validation was demonstrated in 2 studies (Aumente-Maestro et al., 2025; Sobhaninia et al., 2023), which used the 5-fold cross-validation. Just one study combined the two approaches (Maqsood et al., 2022). Most classification studies reported high performance on their own dataset but no evidence of external validation or fine-tuning.

Throughout the reviewed classification studies, 4 studies focused their attention in predicting the primary tumor types investigated, such as glioma, meningioma, and pituitary tumors (Krishnasamy and Ponnusamy, 2023; Maqsood et al., 2022; Rai et al., 2024; Sobhaninia et al., 2023), whereas three papers focused on predicting glioma grading (HGG vs. LGG) (Abd-Ellah et al., 2024; Aumente-Maestro et al., 2025; Zaitoon and Syed, 2023). One study (Mahajan et al., 2023), expanded the classification scope beyond the traditional three classes to include Ependymoma, medulloblastoma, brainstem glioma and pilocytic astrocytoma.

To perform the classification task 32 studies used a ML model. In particular, Maqsood et al. (2022) used a multiclass SVM (Maqsood et al., 2022) to predict the tumor type, reaching on BraTS 2018 dataset an accuracy of 97.47%, 97.22% sensitivity, and 97.94% specificity; and on BTF dataset an accuracy of 98.92%, 98.82% sensitivity, and 99.02% specificity. Aumente-Maestro C. et al. (2025) (Aumente-Maestro et al., 2025) used an XGBoost model to predict the glioma stage (i.e., HGG vs. LGG), in particular the authors trained three different models (i) using only MRI features; (ii) using only segmented features and (iii) the two categories combined. The best model was the third one which achieved the highest accuracy equal to 93.20%, 97.30% sensitivity, and 77.60% specificity; followed by the first model which achieved an accuracy of 91.30%, 96.60% sensitivity, and 70.90% specificity, and lastly the second one showing an accuracy of 81.30%, 91.10% sensitivity, and 43.40% specificity. As XGBoost inherently provides a score for each input feature reflecting its contribution in the prediction, this study was the only one to conduct an explainability analysis showing that the number of voxels of ET, number of voxels of CT, and kurtosis were the most important features.

The remaining studies used DL models to perform both segmentation and classification. Rai et al. (2024) (Rai et al., 2024) using the two-headed UNetEfficientNets showed that B0 achieved a global sensitivity of 97.00%, and accuracy of 94.80%, with tumor-specific sensitivity 92% for meningioma, 99% for glioma, and 100% for pituitary tumors. B1 improved performance further, reaching a global 97.67% sensitivity, and 99.00% accuracy, with meningioma at 94% sensitivity, glioma at 99% sensitivity, and pituitary tumors at 100%. B2 showed slightly lower global sensitivity (i.e., 81.67%) but maintained 93.00% accuracy, with tumor-specific metrics of 67% sensitivity for meningioma, 98% for glioma, and 80% for pituitary tumors. B3 reached a global sensitivity of 98%, and 96% accuracy, with meningioma at 95% sensitivity, glioma at 99% sensitivity, and pituitary tumors at 100%. B4, the best-performing variant, achieved a global sensitivity of 99.00% and 99.40% accuracy, with tumor-specific sensitivity of 98% for meningioma, 99% for glioma, and 100% for pituitary tumors. Finally, B5 showed a global sensitivity of 97.67%, and 99.30% accuracy, with meningioma at 94% sensitivity, glioma at 99%, and pituitary tumors at 100%.

Sobhaninia et al. (2023) (Sobhaninia et al., 2023) used a Multiscale Cascaded Multitask Network, as previously mentioned, created to solve both segmentation and classification tasks. In performing the tumor type classification using the Multiscale Cascaded Multitask Network they achieved a global accuracy of 95.09%, when the aggregation module is included in the model the global accuracy increased reaching an accuracy of 97.99%. Focusing on the single tumor type, the highest precision was reached in classifying glioma (precision=98.32%), followed by meningioma and after pituitary tumor (precision=97.60% and 97.10%, respectively).

Krishnasamy et al. (2023) (Krishna et al., 2025) introduced two hybrid models for brain tumor segmentation and tumor type classification, as stated before. ResNet-50, a deep convolutional neural network, is based on a residual learning framework that mitigates the vanishing gradient problem, enabling the training of very deep networks (He et al., 2016). MobileNet, in contrast, is a lightweight model optimized for mobile and embedded vision applications, with efficiency that makes it particularly suitable for real-time scenarios with limited computational resources (Howard et al., 2017). In terms of performance, Model 1 achieved 94.00% of sensitivity, an accuracy of 98.93%, and specificity at 98.70%. Model 2 showed similar performance, with sensitivity at 92.00%, an accuracy of 98.63%,and specificity at 98.50%.

Lastly, Mahajan et al. (2023) (Mahajan et al., 2023) evaluated the classification of several pediatric brain tumor types, although the specific AI model used was not reported. For brainstem gliomas, the model achieved an accuracy of 94.29%, sensitivity of 88.24%, and specificity of 96.23%.. Ependymomas were classified with 90.00% accuracy, 64.29% sensitivity, and 96.43% specificity. For medulloblastomas, performance reached 98.57% accuracy, 61.90% sensitivity, and 100% specificity. Finally, pilocytic astrocytomas showed 84.29% accuracy, 100% sensitivity, and 78.85% specificity.

Focusing on glioma staging classification, Abd-Ellah et al. (2024) (Abd-Ellah et al., 2024) used a two-pathway with residual-based deep convolutional neural network architecture (TRDCNN), that is composed of four main paths: global, local, merged, and output. The global and local paths independently perform feature extraction through convolutional, ReLU, and max-pooling layers, followed by six residual block stages. Operating in parallel, the two paths are later combined into a merged path comprising batch normalization, ReLU, fully connected, and dropout layers. Finally, the output path includes a softmax classification layer that classified HGG vs. LGG images, reaching an accuracy of 98.88%, a specificity of 99.00%, and a sensitivity of 98.66%.

Zaitoon and Syed (2023) (Zaitoon and Syed, 2023) employed the Deep Brain Tumor Convolutional Neural Network (DBT-CNN) for glioma staging classification using the segmented output from RRU-Net2+. DBT-CNN consists of convolutional, pooling, activation, and fully connected layers, with three convolutional layers followed by layer normalization, ReLU activation, three max-pooling layers, a dropout layer, and a final softmax layer. Classification was performed both on the entire mask and on precisely extracted tumor areas. The model achieved high performance for both tumor types. For HGG, accuracy was 98.51 ± 0.18%, 97.23 ± 0.28% sensitivity, and 98.62 ± 0.46% specificity. For LGG, the accuracy was 99.28 ± 0.18%, 97.83 ± 1.22% sensitivity, 98.88 ± 0.16% specificity.

To summarize, the average accuracy for tumor type classification was 96.95%, ranging from a minimum of 91.79% to a maximum of 99.40%. Regarding glioma staging the average accuracy was equal to 95.97%, with a minimum of 91.30% and a maximum value of 98.90%.

Figure 3 presents a bubble chart illustrating the distribution of model performance, where the x-axis represents sensitivity, the y-axis represents specificity, and the bubble size reflects test size. This Figure highlights that most of the models reported in the selected papers presented either high sensitivity or specificity, and only one paper presented a higher sensitivity and a lower specificity without any influence of the sample size used in the testing phase.

**Figure 3.**
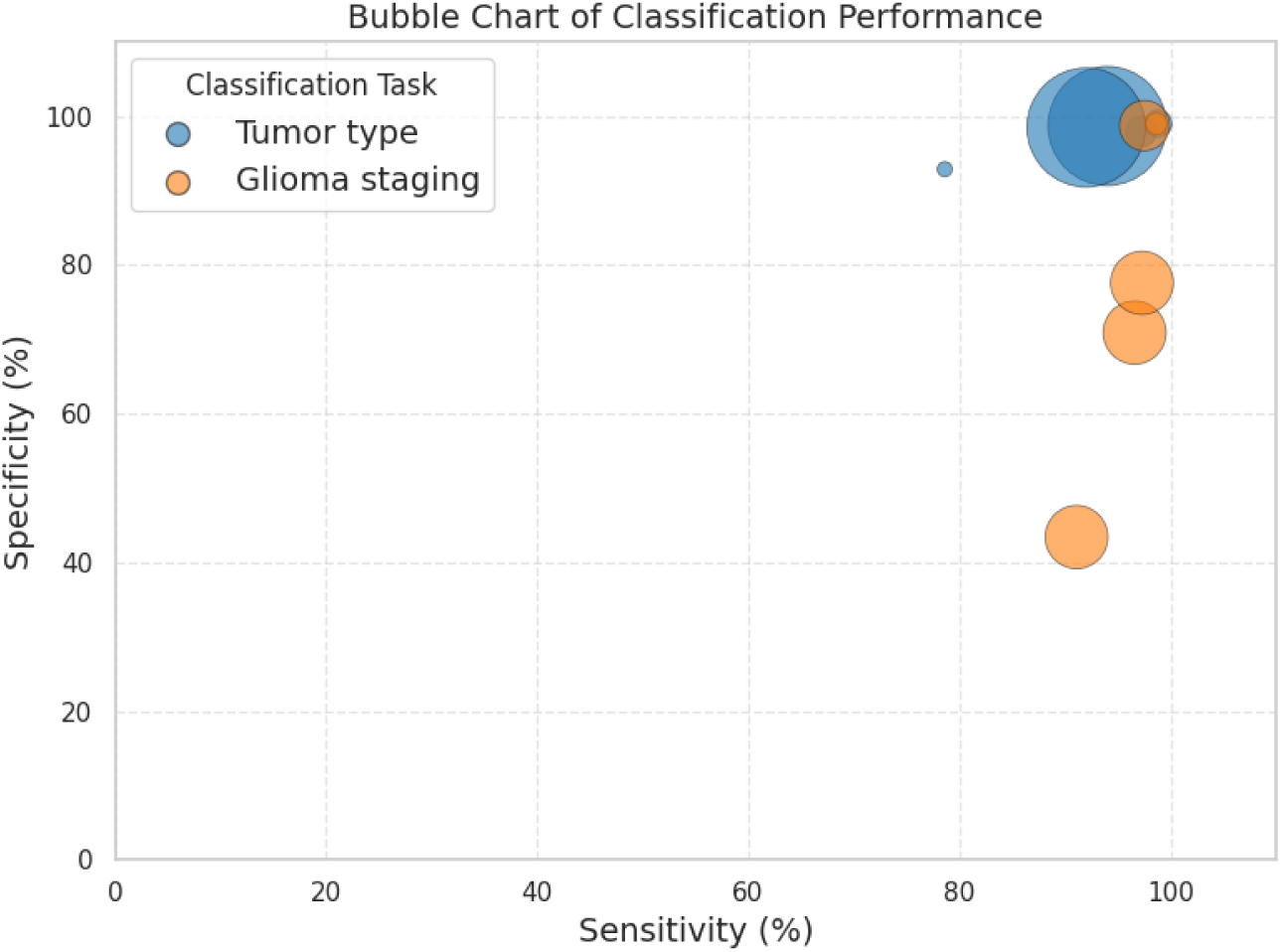
Bubble chart illustrating the distribution of models according to sensitivity (x-axis) and specificity (y-axis), plotted when reported. Each bubble represents a model, with its size proportional to the test sample size and color indicating the classification task. Models located in the upper right corner exhibit both high sensitivity and specificity and thus correspond to the best-performing approaches. Conversely, models in the upper left show high specificity but low sensitivity, while those in the bottom right display high sensitivity but lower specificity. This visualization enables immediate comparison of performance across different classification tasks when these metrics are available.

To facilitate the interpretation of the results, Figure 4 summarizes the complete methodological pipeline adopted by the 8 selected studies for segmentation and classification tasks, from preprocessing to model architecture and performance evaluation. Only methods reported in at least 25% of the included studies were considered.

**Figure 4.**
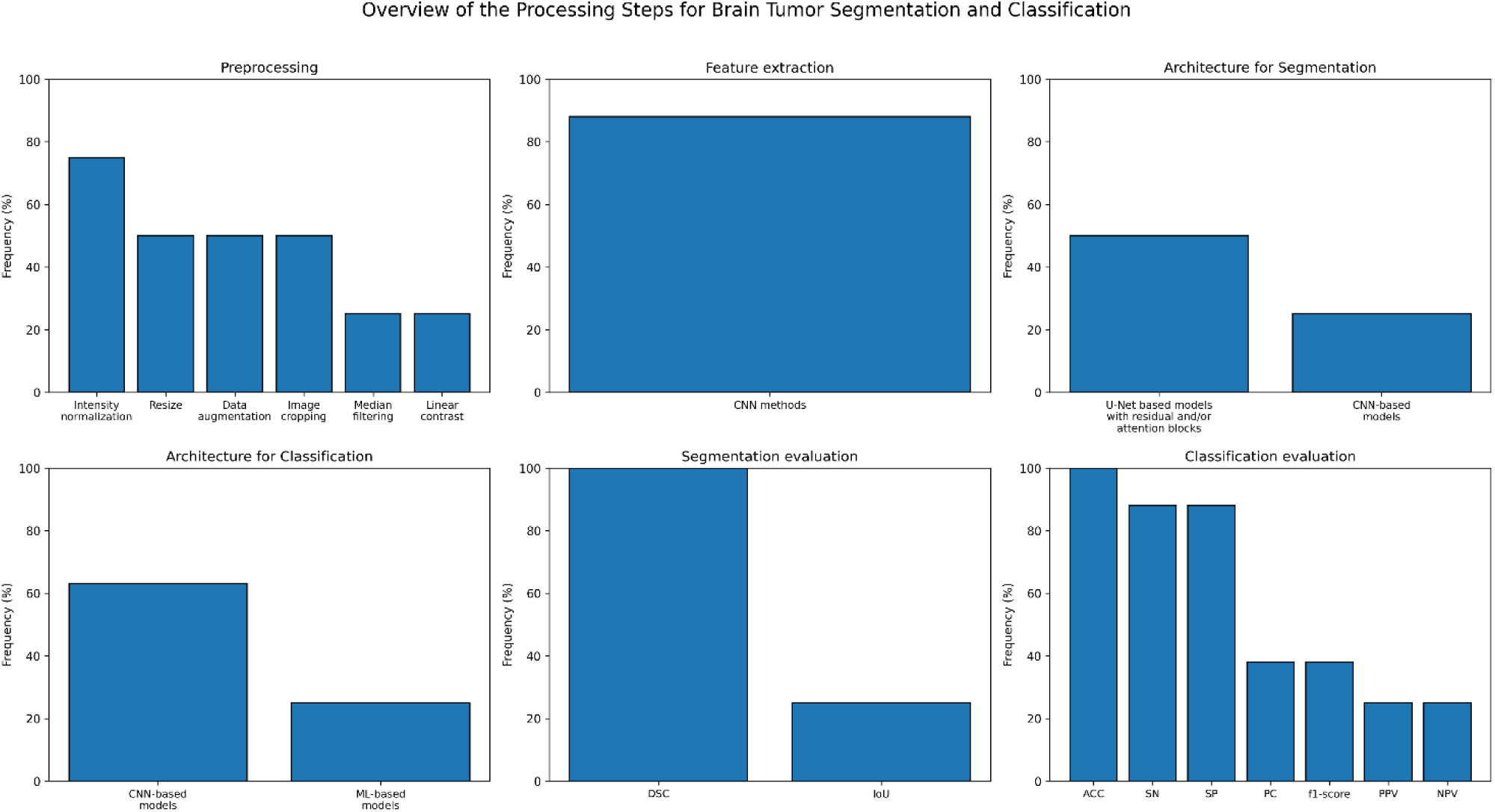
Summary of the methodological pipeline used by studies included in this systematic review that tackled both brain tumor segmentation and classification The relative frequency of approaches adopted for each step is reported. *Legend: CNN: convolutional neural network, ML: machine learning, ACC: accuracy, SN: sensitivity, SP: specificity, PC: precision, PPV: positive predictive value, NPV: negative predictive value*

In the preprocessing phase, 75% of the studies performed intensity normalization, while 50% applied image cropping, resizing, and data augmentation. Linear contrast augmentation and median filtering were each adopted by 25% of the studies. Regarding feature extraction, 88% of the selected studies employed CNN-based methods. For segmentation, 50% of the studies used U-Net–based architectures incorporating residual blocks and/or attention mechanisms and 25% traditional CNN models. For classification, 63% of the studies adopted CNN models, whereas 25% used ML approaches, such as Random Forest and XGBoost. Concerning model evaluation, all studies assessed segmentation performance using the DSC, and 25% also reported the IoU. For classification, all studies reported accuracy, while 88% included sensitivity and specificity, 38% precision and F1-score, and 25% positive predictive value (PPV) and negative predictive value (NPV).

### 3.3 Risk of Bias within Studies

#### 3.3.1 Brain Tumor Segmentation

The risk of bias in the included studies, along with the authors’ comments on the seven QUADAS-2 domains, was evaluated. Figure 5 presents the assessment results across these domains for all studies included in the review. Almost all studies (30 out of 31) showed low concerns regarding applicability, as the patient characteristics, study setting, conduction and interpretation of the index test, and the target condition defined by the reference standard were all aligned with the review question. Only one study was rated as having unclear concern because it did not report the validation method used to train the DL model, making it impossible to determine whether there was any overlap between patients used for training and testing (Mahajan et al., 2023). Looking at the other domains, most of the studies used public available datasets that did not enroll patients in a consecutive or random way. Only five studies used custom-made datasets, as previously mentioned, in which patients were retrospectively recruited, or in one case no information was provided on how they performed patient enrollment (Bouget et al., 2022; Gutsche et al., 2023; Mahajan et al., 2023; Sachdeva et al., 2024; Zanier et al., 2023). Regarding the reference standard domain, twenty-five studies were rated as having low risk of bias, as the reference segmentations were either manually generated by three or more experienced clinicians, produced by trained medical students and subsequently verified by experts, or initially generated by an algorithm and then reviewed by clinicians (Abd-Ellah et al., 2024; Ahsan et al., 2025; Aumente-Maestro et al., 2025; Bouget et al., 2022; Chetty et al., 2022; Esmaeilzadeh Asl et al., 2024; Koteswara Rao Chinnam et al., 2022; Lefkovits et al., 2022; Maqsood et al., 2022; Mazher et al., 2024; Ottom et al., 2022; Rahat et al., 2023; Rai et al., 2024; Raza et al., 2023; Rosa et al., 2024; Roy et al., 2023; Sachdeva et al., 2024; Sobhaninia et al., 2023; Tejashwini et al., 2025; Wang et al., 2022; Xu et al., 2024; Yang et al., 2025; Zafar et al., 2024; Zaitoon and Syed, 2023; Zaman et al., 2024). Four studies were rated as having unclear risk due to a lack of information on the clinicians’ level of experience or because segmentation was performed by fewer than three operators (Krishnasamy and Ponnusamy, 2023; Kundal et al., 2024; Shaikh et al., 2025; Zanier et al., 2023). Two studies were rated as high risk, as the segmentations were conducted by only one experienced neurologist (Gutsche et al., 2023; Mahajan et al., 2023). Lastly, in the flow and timing domain, twenty-one studies were rated as having low risk of bias (Ahsan et al., 2025; Aumente-Maestro et al., 2025; Bouget et al., 2022; Esmaeilzadeh Asl et al., 2024; Koteswara Rao Chinnam et al., 2022; Krishnasamy and Ponnusamy, 2023; Kundal et al., 2024; Mahajan et al., 2023; Maqsood et al., 2022; Mazher et al., 2024; Rahat et al., 2023; Rai et al., 2024; Raza et al., 2023; Rosa et al., 2024; Sobhaninia et al., 2023; Wang et al., 2022; Xu et al., 2024; Zafar et al., 2024; Zaitoon and Syed, 2023; Zaman et al., 2024; Zanier et al., 2023). Five studies were rated as high risk due to the exclusion of some patients from the analysis (Abd-Ellah et al., 2024; Chetty et al., 2022; Gutsche et al., 2023; Ottom et al., 2022; Roy et al., 2023), while another five were rated as unclear risk because the number of patients included in the analysis was not reported (Lefkovits et al., 2022; Sachdeva et al., 2024; Shaikh et al., 2025; Tejashwini et al., 2025; Yang et al., 2025).

**Figure 5.**
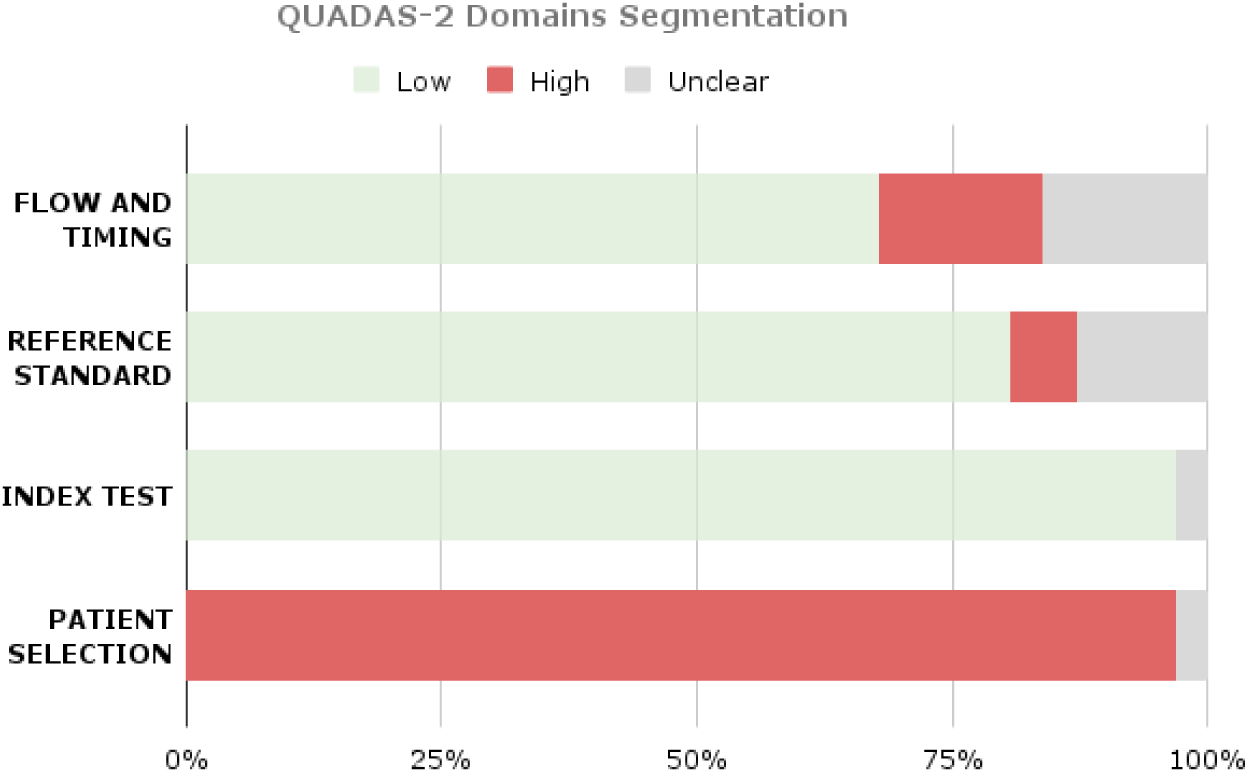
Proportion of included studies categorized by risk of bias assessment (low, high, or unclear).

The risk of bias tools highlighted the following limitations:

- There was no study that performed a consecutive or random recruitment. Only three studies used a custom-made dataset for the analysis (Bouget et al., 2022; Gutsche et al., 2023; Mahajan et al., 2023), but only two studies specified that patients were retrospectively recruited (Bouget et al., 2022; Gutsche et al., 2023);
- Few studies (Abd-Ellah et al., 2024; Chetty et al., 2022; Gutsche et al., 2023; Lefkovits et al., 2022; Roy et al., 2023; Sachdeva et al., 2024; Shaikh et al., 2025; Tejashwini et al., 2025; Yang et al., 2025) did not report information regarding possible inappropriate exclusion of patients, which could introduce selection bias into the analyzed sample;
- The exclusion of patients from the final analysis in a subset of studies (Abd-Ellah et al., 2024; Chetty et al., 2022; Gutsche et al., 2023; Mahajan et al., 2023; Roy et al., 2023) or the lack of information about the final sample (Lefkovits et al., 2022; Sachdeva et al., 2024; Shaikh et al., 2025; Tejashwini et al., 2025; Yang et al., 2025) used for the analysis could lead to attrition bias, affecting the generalizability of results;
- In at least one study (Maqsood et al., 2022), the absence of details on the validation approach used for training the DL model makes it impossible to assess the risk of overfitting and whether the training and testing datasets were truly independent.

#### 3.3.2 Brain Tumor Classification

The risk of bias in the included classification studies was evaluated using the QUADAS-2 framework. Figure 6 summarizes the distribution of judgments across the four domains. Overall, five studies showed low concerns regarding applicability, while three showed high concerns. In the patient selection domain, all studies relied exclusively on publicly available datasets (BTF, BraTS, Kaggle, TCGA) that were not based on consecutive or random patient recruitment, except for one study (Mahajan et al., 2023), resulting in a generally high risk of bias. For the index test domain, most studies were rated as low risk, Classification thresholds were not explicitly reported, and threshold selection procedures were not described (Mahajan et al., 2023)(Maqsood et al., 2022); (Mahajan et al., 2023)(Abd-Ellah et al., 2024; Mahajan et al., 2023), (Abd-Ellah et al., 2024; Mahajan et al., 2023); (Ahsan et al., 2025; Krishnasamy and Ponnusamy, 2023; Maqsood et al., 2022; Rai et al., 2024; Rosa et al., 2024; Shaikh et al., 2025; Sobhaninia et al., 2023; Xu et al., 2024); (Aumente-Maestro et al., 2025; Maqsood et al., 2022; Sobhaninia et al., 2023); (Abd-Ellah et al., 2024; Mahajan et al., 2023)(Aumente-Maestro et al., 2025; Tejashwini et al., 2025) (Abd-Ellah et al., 2024; Mahajan et al., 2023), (Krishnasamy and Ponnusamy, 2023; Mahajan et al., 2023); (Abd-Ellah et al., 2024; Aumente-Maestro et al., 2025; Krishnasamy and Ponnusamy, 2023; Mahajan et al., 2023; Maqsood et al., 2022; Rai et al., 2024; Sobhaninia et al., 2023; Zaitoon and Syed, 2023)(Mahajan et al., 2023). Three studies (Aumente-Maestro et al., 2025; Maqsood et al., 2022; Sobhaninia et al., 2023),(Aumente-Maestro et al., 2025; Maqsood et al., 2022; Sobhaninia et al., 2023) employed robust cross-validation strategies and explicit training, validation, and testing splits and were therefore judged to be at low risk of bias. In the reference standard domain, studies using BraTS and BTF datasets were considered to have low risk of bias ((Aumente-Maestro et al., 2025; Maqsood et al., 2022; Sobhaninia et al., 2023) (Ahsan et al., 2025; Krishnasamy and Ponnusamy, 2023; Maqsood et al., 2022; Rai et al., 2024; Rosa et al., 2024; Shaikh et al., 2025; Sobhaninia et al., 2023; Xu et al., 2024)), In contrast, two studies (Krishnasamy and Ponnusamy, 2023; Mahajan et al., 2023) were judged to have unclear risk. Finally, the flow and timing domain showed mixed results: most studies applied consistent reference standards and clearly reported dataset splits, leading to low-risk judgments, while two studies (Abd-Ellah et al., 2024; Mahajan et al., 2023) were rated as having high risk of bias.

**Figure 6.**
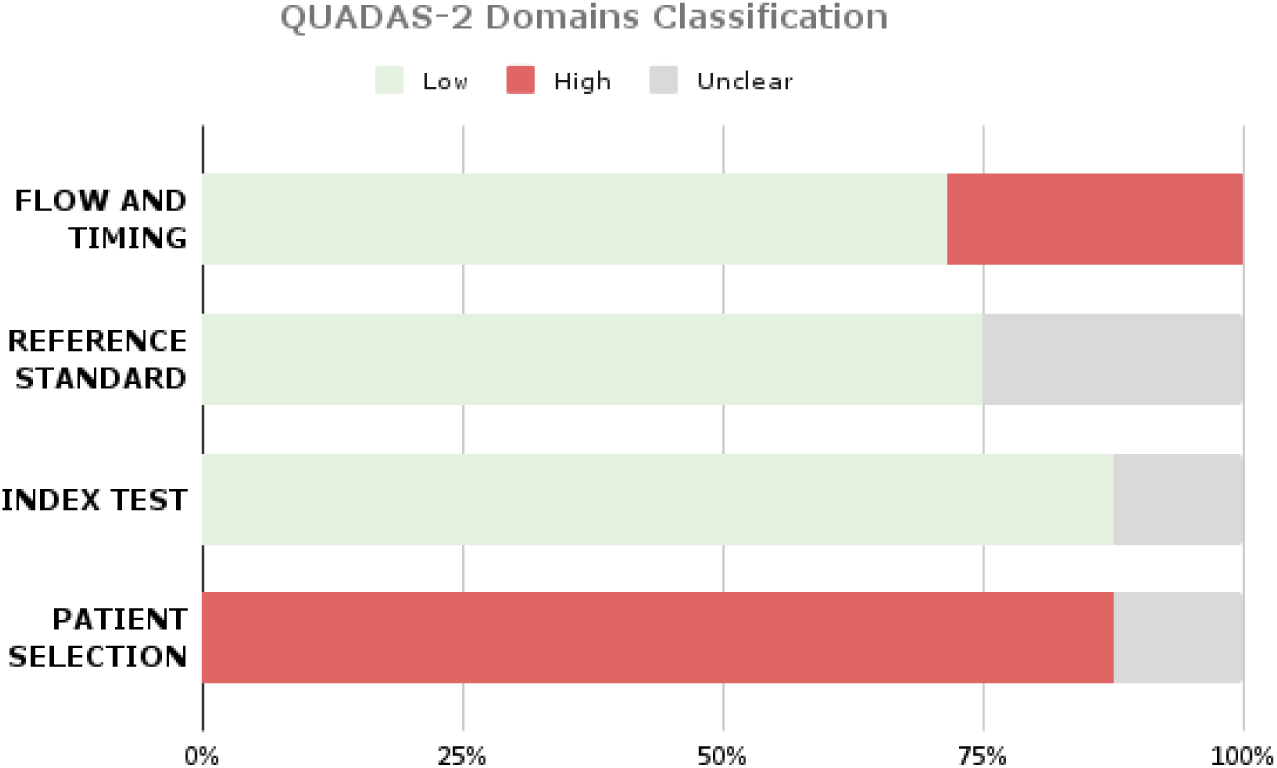
Proportion of included studies that performed both segmentation and classification categorized by risk of bias assessment (low, high, or unclear).

The risk-of-bias analysis highlighted the following limitations:

- Except for one study (Mahajan et al., 2023) that is unclear, no other work employed consecutive or random recruitment, but relied on curated public dataset that led to high risk of bias;
- Thresholds for classification and validation approaches were not prespecified in any study, thus introducing potential bias in index test performance (Mahajan et al., 2023)(Maqsood et al., 2022); (Mahajan et al., 2023)(Abd-Ellah et al., 2024; Mahajan et al., 2023), (Abd-Ellah et al., 2024; Mahajan et al., 2023); (Ahsan et al., 2025; Krishnasamy and Ponnusamy, 2023; Maqsood et al., 2022; Rai et al., 2024; Rosa et al., 2024; Shaikh et al., 2025; Sobhaninia et al., 2023; Xu et al., 2024); (Aumente-Maestro et al., 2025; Maqsood et al., 2022; Sobhaninia et al., 2023); (Abd-Ellah et al., 2024; Mahajan et al., 2023)(Aumente-Maestro et al., 2025; Tejashwini et al., 2025) (Abd-Ellah et al., 2024; Mahajan et al., 2023), (Krishnasamy and Ponnusamy, 2023; Mahajan et al., 2023); (Abd-Ellah et al., 2024; Aumente-Maestro et al., 2025; Krishnasamy and Ponnusamy, 2023; Mahajan et al., 2023; Maqsood et al., 2022; Rai et al., 2024; Sobhaninia et al., 2023; Zaitoon and Syed, 2023)(Mahajan et al., 2023);
- Patient-selection bias was generally high, while it was judged as unclear in one case (Mahajan et al., 2023), due to uncertain sampling methods and inappropriate exclusion criteria;
- Bias related to flow and timing was generally low, although two studies (Abd-Ellah et al., 2024; Mahajan et al., 2023) showed high risk due to incomplete inclusion of all patients or unclear reporting of analysis steps.

## 4. Discussion

The aim of this review was to examine the existing literature on DL-based approaches for the automatic segmentation of gliomas, and, when performed, to analyze the AI methods used for automatic tumor type classification or glioma staging. We reported the results of the selected studies in terms of the DL models employed, the segmented regions and/or predicted classes, and their performance. We further focused on sample characteristics, datasets, imaging modalities, image pre-processing, mask post-processing, feature extraction techniques, and XAI algorithms. For this purpose, we reviewed 31 studies published between 2022 and 2025 that performed glioma segmentation using DL algorithms. Among these, 8 studies investigated either glioma segmentation or classification using AI models trained with interpretable or non-interpretable features as part of the diagnostic workflow.

All the studies included glioma cases, reflecting their prevalence and clinical importance. A smaller subset also included meningioma and pituitary tumor cases. Although meningiomas are relatively easy to segment and pituitary tumors occur more sporadically, gliomas present significant challenges due to their heterogeneous appearance, diffuse and infiltrative growth patterns, and unclear tumor boundaries. These factors make manual delineation subjective and inconsistent (Bakas et al., 2018; Menze et al., 2015). Therefore, accurate segmentation is crucial for treatment planning, surgical navigation, and longitudinal disease monitoring, significantly influencing patient outcomes. This underlines the urgent need for fully automated, objective, and less time-consuming techniques to reduce inter-observer variability and improve clinical workflow efficiency, a necessity strongly emphasized by recent studies (Liu et al., 2023).

Gordillo et al. (2013) (Gordillo et al., 2013) identified several automatic methods for brain tumor segmentation, including AI-based model approaches; however, these require large amounts of training data to perform effectively. A recent review (Magadza and Viriri, 2021), which analyzed the utility of DL models for segmenting various brain tumor types, concluded that the scarcity of large-scale, high-quality medical training datasets remains a primary limitation for the performance and generalizability of many segmentation algorithms.

Consistently, our findings indicate that patient cohort sizes varied widely across studies, ranging from 65 to over 3,000 subjects. Most studies relied on publicly-available datasets such as BraTS, BTF, and TCGA-LGG, while only a small fraction used custom-made datasets. Very few studies implemented DL segmentation to assess its effectiveness in real clinical scenarios, such as evaluating treatment response or tumor reduction after resection. Researchers in this field often prefer publicly available datasets to focus on their experiments, despite not directly addressing clinical applications. Conversely, some researchers emphasized the benefits of developing dedicated repositories, noting that collecting data tailored to specific research objectives prior to experimentation can enhance accuracy, improve model generalization, and yield more unbiased results (Abid and Munir, 2025).

It should be noted that the use of private, custom-made datasets is often associated with relatively small sample sizes and data collected within a single institution or following highly specific clinical protocols. While such datasets may better reflect real clinical scenarios, their limited size and lack of heterogeneity can result in an overestimation of performance, particularly when external validation is not performed (Varoquaux and Cheplygina, 2022).

Conversely, the above-reported widespread use of publicly available datasets, such as BraTS, BTF, or TCGA- and Kaggle-hosted collections, although beneficial for benchmarking and reproducibility, may also introduce systematic biases. Several studies have highlighted that public medical imaging datasets may contain acquisition-specific patterns, preprocessing artifacts, class imbalance, or dataset-specific shortcuts that models can inadvertently exploit, leading to overly optimistic performance estimates and limited generalizability to real-world clinical data (DeGrave et al., 2021; Oakden-Rayner et al., 2020). Such limitations must be taken into account while developing AI models and while evaluating the results of such AI-based systems.

When performing glioma segmentation, the choice of image modality is also crucial. The gold standard for glioma imaging is MRI (Rasool and Bhat, 2025), and this is fully consistent with the evidence gathered in our review. Among the included studies, MRI was by far the most widely used imaging modality for both segmentation and glioma classification tasks. The literature suggests that multiparametric MRI is the most suitable approach, and accordingly, the majority of studies combined T1-weighted, T2-weighted, T1ce-weighted, and FLAIR sequences to leverage complementary information about tumor and surrounding tissue.

Equally critical is image pre-processing, which aims to standardize and enhance MRI data before input into DL models. Common steps include bias field correction, skull stripping, intensity normalization, resize, image cropping, and data augmentation (Litjens et al., 2017; Maharana et al., 2022; Poornachandra and Naveena, 2017). These procedures help reduce scanner-related variability and improve the consistency of the input data, which is particularly important when models are trained on multi-center datasets. In line with the existing literature, our findings confirmed that the most frequently applied pre-processing techniques were intensity normalization, resizing, image cropping, and data augmentation. Collectively, the careful selection of MRI modalities and rigorous preprocessing pipelines play a pivotal role in maximizing the performance and generalizability of DL-based glioma segmentation models. Despite these benefits, preprocessing adds computational steps and potential sources of error. Furthermore, some studies also incorporated post-processing strategies, such as mask smoothing or morphological operations to refine the predicted segmentations.

As stated by literature, in clinical practice it is important to segment diverse tumor regions to obtain different clinically-relevant information (Cariola et al., 2025). Actually, a small subset of the studies focused on segmenting only WT, but the majority focused on multiple glioma subregions, i.e., ET, CT, NCR, NET and ED.

Performance metrics varied across studies, but the DSC was the most commonly reported, followed by Hausdorff Distance (HD), and IoU. Furthermore, our results confirmed the predominance of CNN-based models, in particular U-Net and its derivatives, as the foundational architecture for glioma segmentation, due to its effectiveness in capturing both global contextual information and fine structural details, essential for accurately outlining glioma subregions characterized by irregular borders and heterogeneous intensity patterns. These models demonstrated consistently high performance, as measured by the DSC, with an overall median value of 81%. Among the included studies, these models achieved good performance in the three most segmented regions, with median DSC values of 89% for WT, 84% for CT, and 80% for ET, reflecting their capability to capture both large and small, clinically meaningful tumor areas. The delineation of these challenging regions, along with NCR, NET, and ED, was further enhanced by U-Net frameworks incorporating residual blocks, attention mechanisms, and multi-scale feature aggregation. These architectures effectively captured salient features across multiple MRI modalities, preserved low-level details while integrating high-level contextual information, improved feature learning and boundary delineation, and modeled spatial and channel-wise dependencies, ultimately achieving higher DSCs across all tumor regions. For instance, 3D RR-UNet2+ reached DSCs up to 99.4% in LGG cases across NCR, ED, ET, and NET, while EnsembleUNets obtained 93% DSC for WT, ET, and NET in BraTS 2021. In contrast, simpler models or those not employing multi-scale or attention strategies, such as 2DVNet or ResNet50, showed poor performance (DSC∼0.6–1%), emphasizing the correlation between model residual blocks and attention mechanisms with segmentation accuracy across different tumor regions. Attention-based architectures, such as Attention U-Net and its cascaded variants, effectively suppress irrelevant background information while emphasizing salient tumor regions, allowing the models to focus on clinically relevant structures. These innovations not only enhanced DSC values but also improved the generalizability of models across heterogeneous datasets, underscoring that careful architectural customization remains critical for achieving clinical-grade segmentation performance.

Although more complex models often deliver superior segmentation accuracy, they can be more difficult to interpret and thus less reliable for clinicians. Consequently, explainability analyses, such as those using Grad-CAM, are needed to probe these black-box models and enhance clinical trust (Selvaraju et al., 2016). Nevertheless, only one study in our review applied Grad-CAM to segmentation, demonstrating that the boundary regions of the tumor were the critical areas guiding the model’s prediction. This finding simultaneously underscores the general lack of systematic explainability analyses in current deep learning–based glioma segmentation studies.

Delving into tumour classification, this represents the final and crucial stage of the tumour diagnosis pipeline, as it not only enables differentiation between tumour types or grades but also informs prognosis, treatment planning, and potential therapeutic strategies. Accurate and early classification is therefore essential to translate imaging findings into meaningful clinical decisions. The reviewed studies addressing tumour classification reported overall high performance, with average accuracies of 96.95% for tumour type identification. Most models relied on CNNs for feature extraction, often embedded within segmentation frameworks to jointly optimize both tasks and capture hierarchical spatial representations directly from MRI data. In tumour-type classification studies, CNN-based models achieved accuracies ranging from 91.79% to 99.40%, demonstrating strong discriminative power in distinguishing among glioma, meningioma, and pituitary tumours. It is worth noting that in only one case such models were combined with clinical and radiological features. Despite these promising results, the limited use of external validation and the prevalence of hold-out strategies raise concerns about model generalizability.

Regarding glioma staging, only a few studies specifically focused on discriminating between HGG and LGG, achieving accuracies between 91.30% and 98.90%. These models employed CNN or hybrid deep learning architectures, whereas one study uniquely combined radiomic and segmentation-derived features within an XGBoost framework, performing an explainability analysis that identified the number of tumour voxels and intensity-based metrics as key predictors.

Also in this context, explainability emerges as a pivotal component for advancing the clinical applicability of AI-based tumour classification. Beyond improving transparency, explainable models can reveal which imaging features strongly drive classification decisions, allowing clinicians to verify the biological plausibility of the results and to better trust model outputs. This interpretive layer is especially relevant in neuro-oncology, where treatment decisions are high-stakes and errors may have serious consequences. Explainable frameworks can also help uncover novel imaging biomarkers and support hypothesis generation by linking learned representations to known pathophysiological mechanisms. Despite its clear potential, explainability remains underrepresented in current literature also when considering the tumour-classification task, with only 1 out of 8 studies incorporating post-hoc analyses to visualize feature importance.

Although these models achieved excellent results, the lack of standardized MRI protocols, dataset heterogeneity, and limited external validation constrain comparability and robustness across studies. Moreover, several methodological limitations in the included studies were identified. Most studies did not provide sufficient details regarding patient selection or potential exclusion criteria, a limitation largely attributable to the reliance on publicly available datasets. In the segmentation studies, reference standards were generally appropriate; however, a few studies failed to specify the number and expertise of annotators or the procedures adopted for annotation. To assess study quality, the QUADAS tool was applied, though it should be acknowledged that certain items are not fully suited to machine learning–based studies. In this field, biases often arise during data preparation, for instance, when training and testing data are not properly separated, leading to optimistic performance evaluation due to overfitting and consequently poor generalization to unseen data. Additionally, in the classification studies, the QUADAS framework identified additional issues regarding the reference standard and decision thresholds. In many cases, tumor classifications were based on inconsistent diagnostic criteria or insufficient histopathological confirmation, which may compromise the reliability of the established ground truth.

This systematic review also highlights the following limitations that may affect the generalizability and clinical applicability of current models. First, the number of included studies remains relatively limited compared to the total volume of research available in the field. Although the automated segmentation of gliomas using DL algorithms is a rapidly expanding area, many studies still focus primarily on tumor detection rather than precise segmentation. Moreover, when segmentation is performed, appropriate performance metrics such as the DSC or the IoU, which more accurately quantify the spatial overlap between the reference standard and the predicted segmentation mask, are not consistently reported. The lack of standardized evaluation metrics complicates cross-study comparison and hinders the objective assessment of model performance. Furthermore, the majority of the included studies relied on publicly-available datasets, which do not follow consecutive or random patient recruitment, potentially introducing selection bias. External validation on independent cohorts was limited, and only a few studies evaluated models on diverse or multicentric data, raising concerns regarding real-world applicability. Variability in imaging protocols, pre-processing techniques, and post-processing strategies further complicates direct comparison across studies. The diversity of performance metrics employed, including DSC, IoU, and accuracy, also hinders quantitative comparison across studies. The high computational requirements for complex DL architectures may restrict the reproducibility and implementation of these methods in resource-limited clinical settings. Finally, there is a clear lack in the application of explainability analysis for both segmentation and classification, to better understand AI model reasoning and make it more trustworthy for clinicians.

Future research should therefore focus on integrating multiparametric MRI data, adopting standardized imaging protocols, and implementing rigorous cross-validation and external testing. Furthermore, the systematic incorporation of explainability frameworks is essential to enhance reproducibility, trustworthiness, and ultimately, the clinical applicability of AI-based tumour segmentation and classification systems.

## Data availability

No data were used for the research described in the article.

## CRediT authorship contribution statement

**Simona Aresta:** Data curation, Formal analysis, Writing - original draft, Writing – review and editing. **Cinzia Palmirotta:** Data curation, Formal analysis, Writing - original draft. **Muhammad Asim:** Data curation, Formal analysis, Writing - original draft. **Petronilla Battista:** Writing – review and editing. **Claudia Cava:** Writing – review and editing. **Pietro Fiore:** Writing – review and editing. **Andrea Santamato:** Writing – review and editing. **Paolo Vitali:** Writing – review and editing. **Isabella Castiglioni:** Writing – review and editing. **Gennaro D’Anna:** Writing – review and editing. **Leonardo Rundo:** Writing – review and editing. **Christian Salvatore:** Conceptualization, Supervision, Methodology, Writing – review and editing.

